# Safety and efficacy of the blood-stage malaria vaccine RH5.1/Matrix-M in Burkina Faso: interim results of a double-blind, randomised, controlled phase 2b trial in children

**DOI:** 10.1101/2024.10.15.24315473

**Authors:** Hamtandi M. Natama, Jo Salkeld, Athanase Somé, Seyi Soremekun, Salou Diallo, Ousmane Traoré, Toussaint Rouamba, Florence Ouédraogo, Edouard Ouédraogo, K. Carine Sonia Daboné, Nadine A. Koné, Z. Michael John Compaoré, Miguel Kafando, Massa dit Achille Bonko, Fabé Konaté, Hermann Sorgho, Carolyn M. Nielsen, Dimitra Pipini, Ababacar Diouf, Lloyd D. W. King, Umesh Shaligram, Carole A. Long, Jee-Sun Cho, Alison M. Lawrie, Katherine Skinner, Rachel Roberts, Kazutoyo Miura, John Bradley, Sarah E. Silk, Simon J. Draper, Halidou Tinto, Angela M. Minassian

## Abstract

**Background:** Two pre-erythrocytic vaccines (R21/Matrix-M and RTS,S/AS01) are now approved for *P. falciparum* malaria. However, neither induces blood-stage immunity against parasites that breakthrough from the liver. RH5.1/Matrix-M, a blood-stage *P. falciparum* malaria vaccine candidate, was highly immunogenic in Tanzanian adults and children. We therefore assessed the safety and efficacy of RH5.1/Matrix-M in Burkinabe children.

**Methods:** In this double-blind, randomised, controlled phase 2b trial, RH5.1/Matrix-M was given to children aged 5-17 months in Nanoro, Burkina Faso – a seasonal malaria transmission setting. Children received either three intramuscular vaccinations with 10 µg RH5.1 protein with 50 µg Matrix-M adjuvant or three doses of rabies control vaccine, Rabivax-S, given either in a delayed third dose (0-1-5-month) regimen (first cohort) or a 0-1-2-month regimen (second cohort). Vaccinations were completed part-way through the malaria season. Children were randomly assigned 2:1 within each cohort to receive RH5.1/Matrix-M or Rabivax-S. Participants were assigned according to a random allocation list generated by an independent statistician using block randomisation with variable block sizes. Participants, their families, and the study teams were masked to group allocation; only pharmacists who prepared the vaccines were unmasked.

Vaccine safety, immunogenicity, and efficacy were evaluated. Co-primary objectives assessed were: i) safety and reactogenicity of RH5.1/Matrix-M, and ii) protective efficacy of RH5.1/Matrix-M against clinical malaria from 14 days to 6 months post-third vaccination in the per-protocol population. This trial is registered with ClinicalTrials.gov (NCT05790889).

**Findings:** From 6^th^ to 13^th^ April and 3^rd^ to 7^th^ July 2023, 412 children aged 5-17 months were screened, and 51 were excluded. A total of 361 children were enrolled in this study. In the first cohort, 119 were assigned to the RH5.1/Matrix-M delayed third dose group, and 62 to the equivalent rabies control group. The second cohort included 120 children in the monthly RH5.1/Matrix-M group and 60 in the equivalent rabies control group. The final vaccination was administered to all groups from 4^th^ to 21^st^ September 2023. RH5.1/Matrix-M in both cohorts had a favourable safety profile and was well tolerated. Most adverse events were mild, with the most common being local swelling and fever. No serious adverse events were reported. A Cox regression model was used to analyse the primary endpoint of time to first episode of clinical malaria, according to the primary case definition, within 14 days to 6 months post-third vaccination. Comparing the RH5.1/Matrix-M delayed third dose regimen with the pooled control groups resulted in vaccine efficacy of 55% (95% CI 20-75%; p=0·0071). The same analysis showed a vaccine efficacy of 40% (95% CI -3-65%; p=0·066) when comparing the monthly regimen with the pooled control groups. Participants vaccinated with RH5.1/Matrix-M in both cohorts showed high concentrations of anti-RH5.1 serum IgG antibodies 14 days post-third vaccination, and the purified IgG showed high levels of *in vitro* growth inhibition activity (GIA) against *P. falciparum*; these responses were higher in RH5.1/Matrix-M vaccinees who received the delayed third dose, as opposed to monthly, regimen.

**Interpretation:** RH5.1/Matrix-M appears safe and highly immunogenic in African children and shows promising efficacy against clinical malaria when given in a delayed third dose regimen. This trial remains ongoing to further monitor efficacy over time.

**Funding:** The European and Developing Countries Clinical Trials Partnership; the UK Medical Research Council; the National Institute for Health and Care Research Oxford Biomedical Research Centre; the Division of Intramural Research, National Institute of Allergy and Infectious Diseases; the US Agency for International Development; and the Wellcome Trust.

## Introduction

Malaria caused by the *Plasmodium falciparum* parasite continues to exert a heavy disease burden across sub-Saharan Africa.^1^ However, two first-generation partially-effective pre-erythrocytic vaccines (RTS,S/AS01 and R21/Matrix-M) are now recommended for malaria prevention in children using a 4- dose schedule from around 5 months of age. These two vaccines are similar in design and target the liver- invasive sporozoite.^2,3^ However when this immunity fails or wanes over time, and sporozoites infect the liver, blood-stage infection ensues with risk of clinical disease. Vaccination against the blood-stage merozoite would thus provide a second line of defence. However, development of an effective blood-stage vaccine has proved challenging,^4^ with all prior phase 2b field efficacy trials reporting either no or minimal efficacy, or evidence of strain-specific efficacy linked to target antigen polymorphism.^5–9^

Identification of the reticulocyte-binding protein homologue 5 (RH5) as a vaccine target,^10^ has since transformed the blood-stage *P. falciparum* vaccine field. This merozoite protein forms an essential interaction with basigin/CD147 on the human red blood cell during invasion^11^ and, unlike previous antigen targets, is almost completely conserved, likely explaining the human-species tropism of this parasite.^12^ We have previously demonstrated high-level efficacy of RH5-based vaccination in non-human primates (NHPs)^13^ and a significant reduction in parasite growth rate in UK adult vaccinees post- challenge.^14^ In both the NHP and human studies we observed a strong correlation between *in vivo* reduction of parasite growth and *in vitro* growth inhibition activity (GIA),^13,14^ since validated as a mechanistic immune correlate in NHPs.^15^ We have also reported promising safety and reactogenicity data from four phase 1a/b trials of RH5-based vaccine candidates in 193 adults, children and infants in the UK and Tanzania.^14,16–18^ Notably, the RH5.1 protein^19^ with Matrix-M adjuvant, in 5-17 month-old Tanzanian children, gave the highest levels of human vaccine-induced GIA ever reported, exceeding the protective threshold identified in the NHP model.^13,18^ We therefore initiated a phase 2b trial (called “VAC091”) of this vaccine candidate in children aged 5-17 months in Nanoro, a seasonal malaria transmission setting in Burkina Faso, to assess its protective efficacy against clinical malaria.

## Methods

### Study design and participants

VAC091 is a double-blind, randomised, controlled phase 2b trial conducted by the Institut de Recherche en Sciences de la Santé at the Clinical Research Unit of Nanoro, Burkina Faso and sponsored by the University of Oxford. Participants aged 5-17 months were recruited at the Siglé trial site, located within the Nanoro Health and Demographic Surveillance System (HDSS) catchment area. This covers 24 villages, with a population of over 63,000 inhabitants. Nanoro is an area where malaria transmission occurs throughout the year, but with a marked peak during the rainy season (June to November).

Eligible participants were recruited into four groups (figure 1; appendix p 24). Groups 1 and 2 (first cohort) received the delayed third-dose (0-1-5-month) regimen (“delayed regimen”) of rabies or RH5.1/Matrix-M vaccines, respectively. Groups 3 and 4 (second cohort) received these vaccines in a “monthly regimen” (0-1-2-month). The third vaccination was given simultaneously across all four groups and completed part-way through the malaria season. Field workers collected data on indoor residual spraying (IRS) of households, insecticide treated net (ITN) use and if the nets were adequate (according to if holes were present), and number of doses and months of seasonal malaria chemoprevention (SMC) taken by the participant during the malaria transmission season.

**Figure 1:**
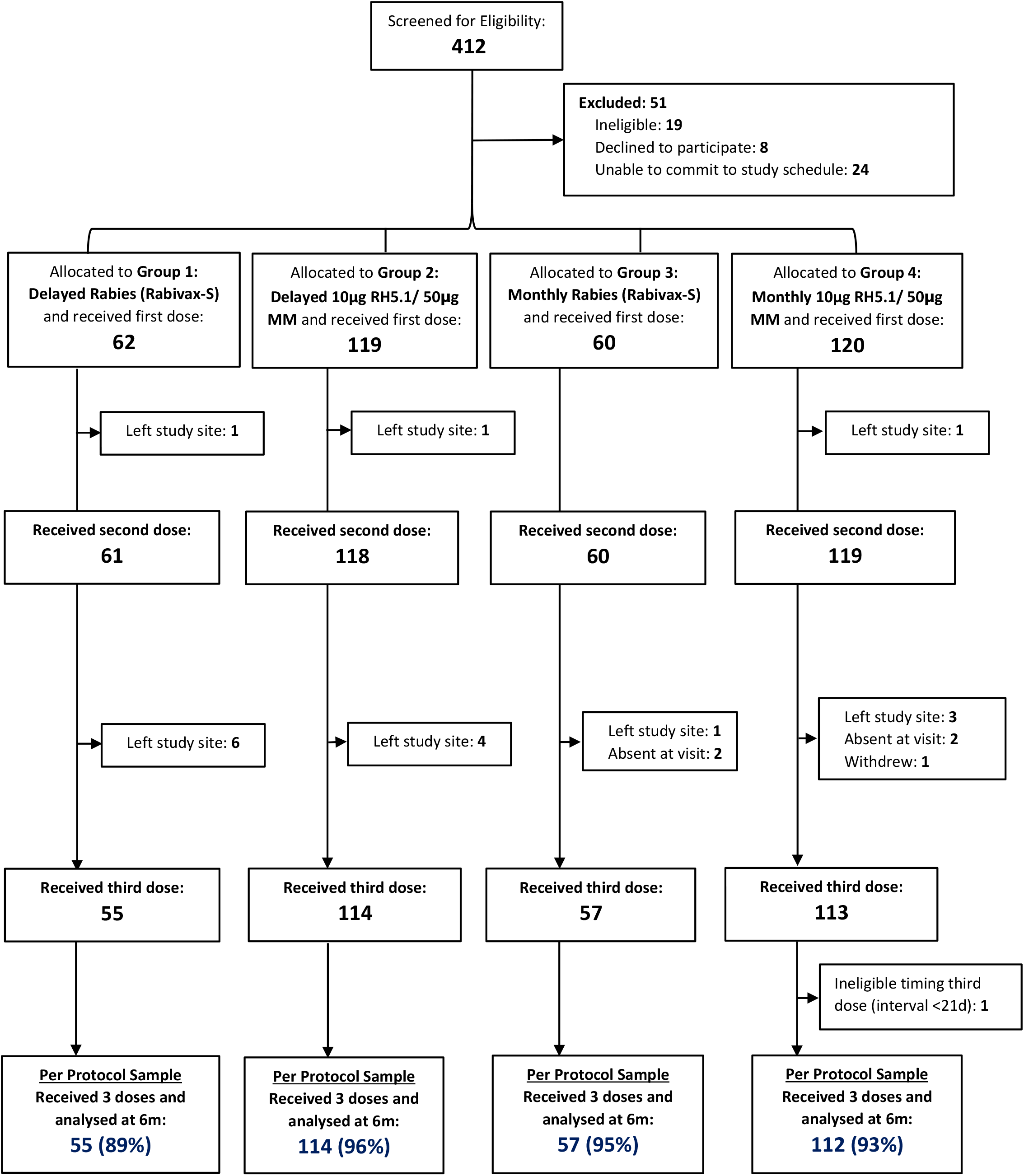
Trial profile.

After community sensitisation, a list of eligible children was drawn from the HDSS database, and parents or legally authorised guardians who expressed interest were invited to screening visits. Prior to the recruitment, parents or guardians of participants provided written or thumb-printed consent, which was verbally checked at each study visit. Inclusion criteria specified that participants should be aged 5-17 months at enrolment and aim to be living in the study area for the whole trial duration. Exclusion criteria included significant co-morbidities and participation in other malaria intervention studies and clinical trials. Further details are given in the appendix (pp 8-9).

The trial was approved by the National Ethical Committee of the Ministry of Health (Comité d’Ethique pour la Recherche en Santé, reference number 2022-12-256), and the national regulatory authority, Agence National de Régulation Pharmaceutique (reference 2023/0208/MSHP/SG/ANRP/DHEC/MIK), in Burkina Faso. Ethical approval was granted in the UK by the Oxford Tropical Research Ethics Committee (reference 3-23). This study is registered with ClinicalTrials.gov, NCT05790889.

### Randomisation and masking

Children aged 5-17 months were randomly assigned (1:2) to Groups 1 and 2 (delayed third-dose regimen) in the first cohort and similarly to Groups 3 and 4 (monthly regimen) in the second cohort. A statistician generated a random allocation list, using block randomisation with variable block sizes, and prepared and sealed the envelopes using this list, which was then given to the pharmacist to assign to participants. Both malaria and control vaccines were prepared by the pharmacist using the same type of syringe, and the contents of the syringe were covered with an opaque label. The trial was double-blinded; participants, their families, the central and local study teams and laboratory teams, were masked to group allocation.

Only the pharmacists preparing the vaccines and statisticians were unmasked to group allocation.

### Procedures

The RH5.1 soluble protein was originally produced according to Good Manufacturing Practice (GMP) by the Clinical Biomanufacturing Facility in Oxford, UK.^19^ A second batch was filled in 2021 under GMP by a Contract Manufacturing Organisation in the UK and was used in this trial. A 10µg dose of RH5.1 protein was mixed with 50μg Matrix-M, a potent, saponin-based adjuvant manufactured by Novavax AB (Uppsala, Sweden) and the Serum Institute of India Pvt. Ltd. (SIIPL), immediately before administration. A rabies vaccine (Rabivax-S), manufactured by SIIPL, was the control vaccine. All vaccines were administered intramuscularly into the thigh.

On the day of enrolment, a blood film was performed to check for *Plasmodium* spp. parasites. In the absence of a fever ≥37·5°C and/or history of fever within the last 24 hours, participants proceeded to vaccination, but if then found to be film-positive, they received treatment for malaria in accordance with national guidelines. For each subsequent vaccination, participants were tested for malaria if they had a fever of ≥37·5°C and/or history of fever within the last 24 hours. If their blood film was positive, they were treated for malaria before being vaccinated upon recovery. After each vaccination, local and systemic solicited adverse events (AEs) were collected for 7 days. Unsolicited AEs were collected for 28 days after vaccinations and classified according to MedDRA (version 27·0). Severity and causality of AEs were assessed using standardised methods (appendix pp 10-12, 25-26) and followed up until resolution. Safety laboratory values were measured at 14 days post-second vaccination, day of third vaccination (Groups 1 and 2 only) and two and six months post-third vaccination to look for deviations from baseline. Serious adverse events (SAEs) are being recorded for the whole duration of the study. A Data Safety Monitoring Board (DSMB) review was held after the first vaccination of the first 100 participants.

Parents of participants were advised to attend the trial site or community health centres if their child had any illness or fever, for review and assessment for malaria. After the third vaccination, participants were also visited by field workers approximately every 30 days until 6 months after the third vaccination and a blood spot was taken for parasite quantification and genotyping. If they had a temperature of ≥37·5°C and/or history of fever within the last 24 hours, blood sampling was also performed for blood film microscopy to detect *Plasmodium* spp.

Anti-RH5.1 serum total IgG responses were measured by ELISA against full-length RH5 protein (RH5.1), using standardised methodology as previously described.^18^ Standardised GIA assays were performed by the GIA Reference Center, NIH, USA, using previously described methodology^20^ (appendix p 22-23).

### Outcomes

The co-primary objectives assessed were: i) safety and reactogenicity of RH5.1/Matrix-M, and ii) protective efficacy of RH5.1/Matrix-M against clinical malaria from 14 days to 6 months post-third vaccination. The primary case definition of clinical malaria was presence of an axillary temperature of ≥37·5°C and/or history of fever in the past 24 hours, and *P. falciparum* asexual parasite density >5000 parasites per μL. Secondary case definitions were presence of an axillary temperature of ≥37·5°C and/or history of fever during the last 24 hours, and i) *P. falciparum* parasite density of >0 parasites per μL; or ii) parasite density of >20,000 parasites per μL. Post-hoc analyses assessed additional case definitions with parasite densities of >50,000 and >100,000 parasites per μL. Secondary objectives assessed i) protective efficacy of RH5.1/Matrix-M against clinical malaria from 14 days to 3 months post-third vaccination; ii) protective efficacy of RH5.1/Matrix-M against prevalent moderate or severe anaemia at 6 months post-third vaccination; and iii) the humoral immunogenicity of RH5.1/Matrix-M. Analysis of other pre-specified secondary objectives regarding the primary vaccination series remain in progress and will be reported after the end of the trial (appendix pp 12-15).

## Statistical analysis

It was estimated that 104 children per arm would give 90% power to detect a 50% vaccine efficacy in either Group 2 or 4 compared to the pooled controls (Groups 1 and 3) if there were 1·2 episodes of clinical malaria per child in the first 6 months of follow up in the control arm (appendix p 7). The rate of 1·2 events per child comes from a study previously conducted in the same area.^3^ 120 children per arm were recruited to allow for 15% loss to follow-up.

For the co-primary endpoint of vaccine safety and reactogenicity, odds ratios comparing the proportion of doses that resulted in solicited AEs were calculated using logistic regression. For the co-primary endpoint of vaccine efficacy, Cox regression models were used to analyse time to first episode of clinical malaria (as per the primary case definition) within 6 months post-third vaccination. Follow up time started 14 days post-third vaccination. Vaccine efficacy (VE) was calculated as 1 minus the hazard ratio (HR). The secondary and additional case definitions of clinical malaria were analysed in the same way. A secondary analysis of VE against all clinical malaria episodes was also carried out, using Cox regression models with a robust standard error to account for multiple episodes. Episodes occurring within 14 days of a previous episode were classed as the same event. For participants without an episode of clinical malaria, their time was censored at the date of their withdrawal or the date of their 6-month blood sampling (noting no deaths have occurred in this trial). A secondary analysis of time to first episode of clinical malaria (analysed as per the primary endpoint) but restricted to episodes occurring within 3 months of the third vaccination was also carried out. The primary comparisons were pre-specified as being between Group 2 and the pooled control Groups 1 and 3, and between Group 4 and the pooled Groups 1 and 3, with comparison of Groups 1 to 2 and 3 to 4 as a supplementary analysis. A secondary analysis adjusted for confounding factors including total number of rounds of SMC received, ITN use (adequate or not) the night before the screening visit, and age at randomisation (5-8 months, 9-12 months, or 13-17 months).

### Event rates of malaria are also reported here, but for information only

The primary analysis of VE was based on a per-protocol population, which included all participants who received three vaccinations correctly and within the pre-specified time period. Secondary analyses included the intention-to-treat population of any child who received at least the first dose of vaccine and remained in the site at the start of the follow-up period (2 weeks post their third dose).

Assays to measure *in vitro P. falciparum* GIA and serum anti-RH5.1 IgG responses were conducted on blood samples taken at baseline (screening) and at day 14 post-third vaccination. GIA data were expressed as percentages and compared between the combined control groups and each RH5.1/Matrix-M vaccine group, and between the two RH5.1/Matrix-M vaccine groups. The measure of effect was the difference in mean percentage GIA, with inference done using the bootstrap method. Serum anti-RH5.1 IgG concentrations measured by ELISA were log10-transformed and the same between-arm comparisons performed by linear regression.

All statistical analyses were performed by independent statisticians using Stata, version 18.

## Results

From 6^th^ to 13^th^ April 2023, 208 children aged 5-17 months were screened, and from 3^rd^ to 7^th^ July 2023, a further 204 children aged 5-17 months were screened (figure 1). Fifty-one were excluded and 361 enrolled across two cohorts. In the first cohort, 119 children were allocated to receive RH5.1/Matrix-M in a delayed third-dose regimen (Group 2) and 62 children allocated to rabies vaccination in the same regimen (Group 1). In the second cohort, 120 children were allocated to RH5.1/Matrix-M in a monthly regimen (Group 4) and 60 children allocated to receive rabies vaccination in the same regimen (Group 3). The final vaccination of the primary series was administered contemporaneously across both cohorts from 4^th^ to 21^st^ September 2023. Twenty-two participants received fewer than three vaccinations, and one participant in Group 3 who received three vaccinations had an interval of less than 21 days between the second and third vaccine doses, so 338 participants were included in the per-protocol analysis at six months post-third vaccination.

Baseline characteristics were similar across the four study groups, with the overall mean age at screening of 10·5 months (SD 3·6), and 177 (52·4%) of the enrolled participants being female (appendix p 31). Of the 361 participants, 290 (85·8%) slept under an adequate ITN the night before screening, and 333 (98·5%) received at least one round of SMC. However, only 8 (2·4%) of 361 participants lived in a house which had received IRS with insecticide in the past year.

Analysis of the primary endpoint of time to first episode of clinical malaria (as per the primary case definition) within 14 days to 6 months post-third vaccination, using a Cox regression model to compare the RH5.1/Matrix-M delayed third dose regimen with the pooled control groups, resulted in VE of 55% (95% CI 20-75%; p=0·0071); the same analysis showed a VE of 40% (95% CI -3-65%; p=0·066) when comparing the monthly regimen with the pooled control groups (table 1 and figure 2A).

**Figure 2:**
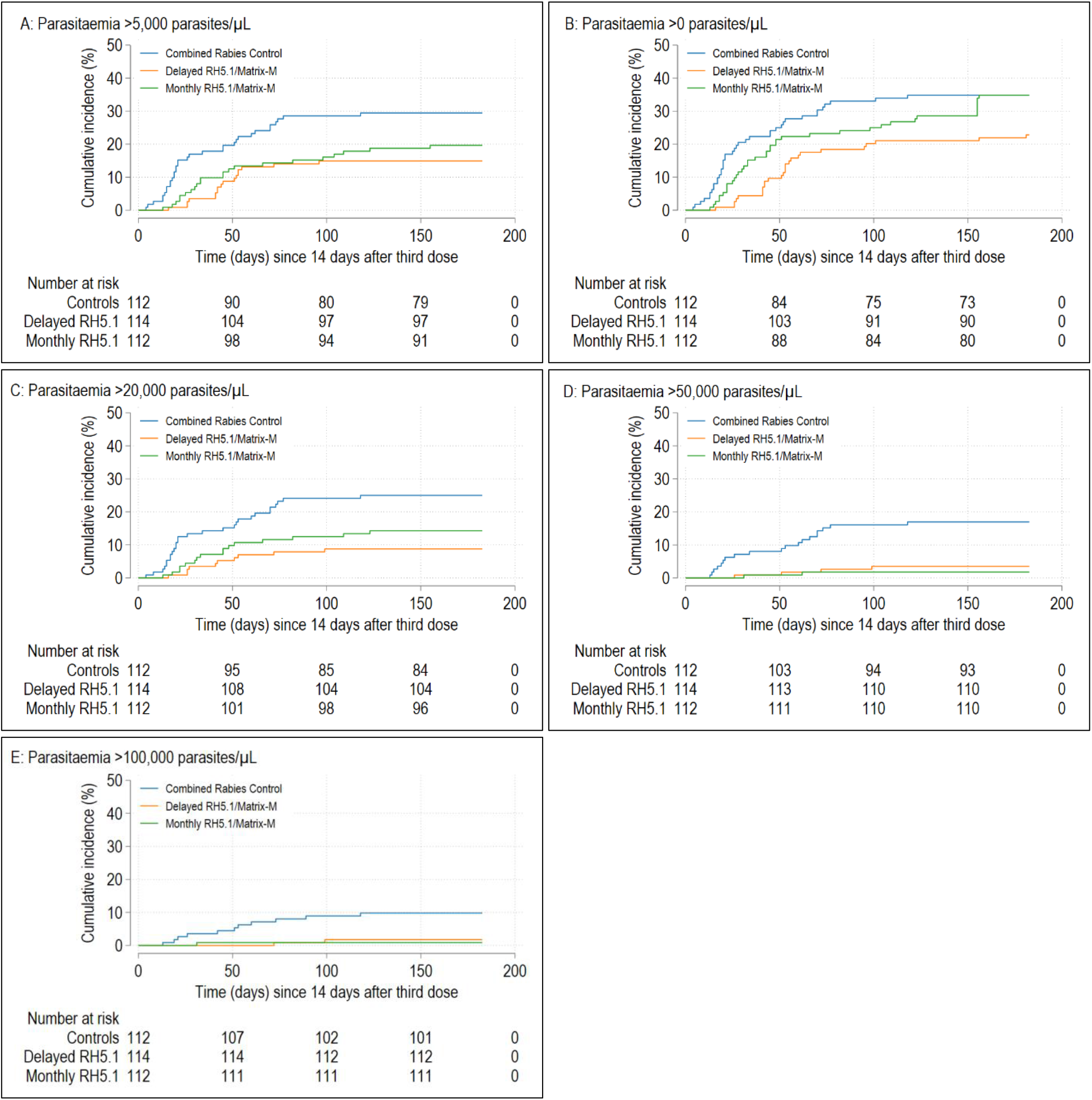
Kaplan-Meier curves showing time to first episode of clinical malaria from 14 days to 6 months post-third vaccination.

**Table 1:**
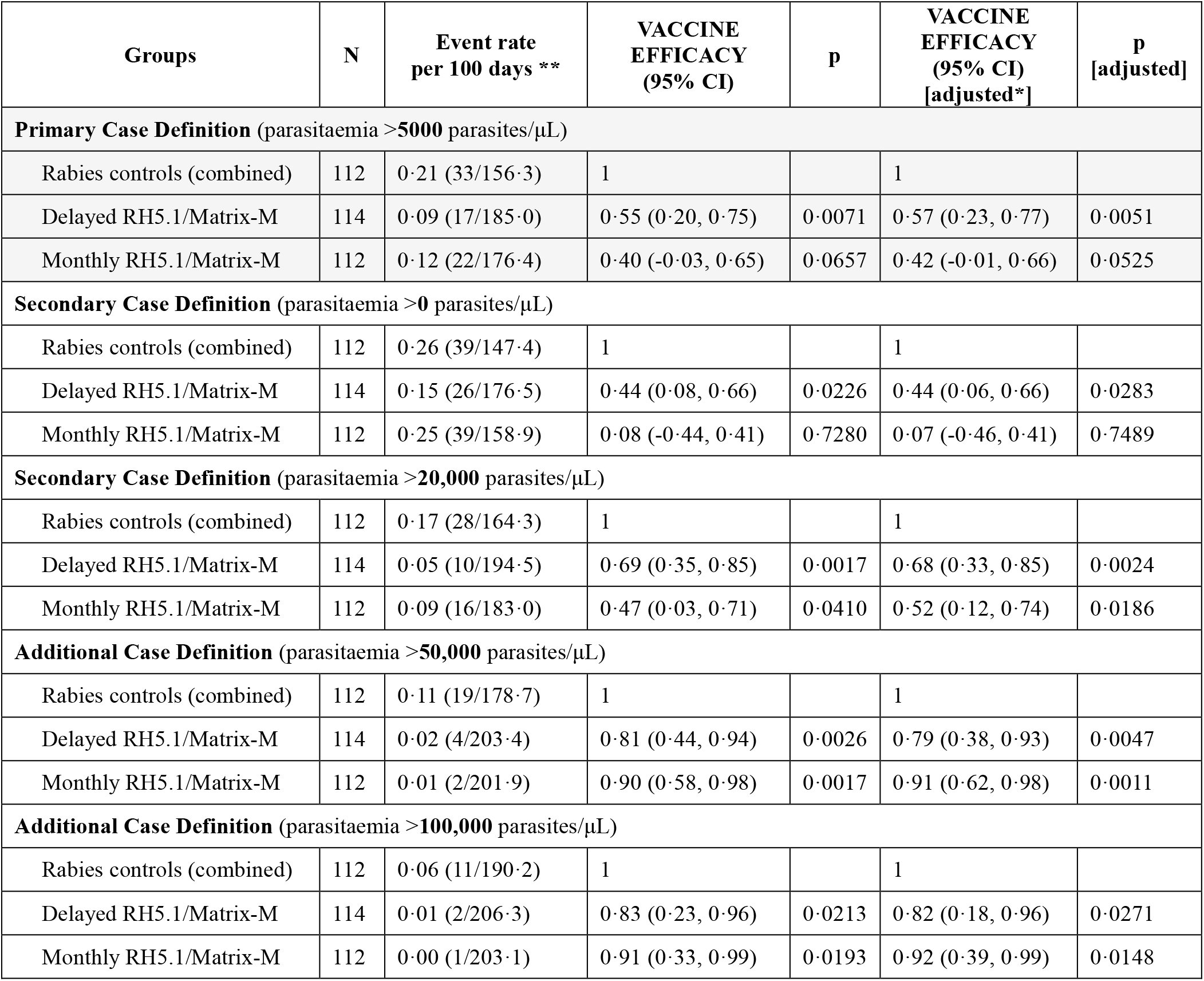
Analysis of time to first episode of clinical malaria from 14 days to 6 months post-third vaccination.

During this primary objective study period (within 14 days to 6 months post-third vaccination) a first episode of clinical malaria occurred in 17 of 114 participants (14·9%) in the delayed RH5.1/Matrix-M group (event rate 0·09/100 child days); 22 of 112 participants (19·6%) in the monthly RH5.1/Matrix-M group (event rate 0·12/100 child days); and 33 of 112 participants (29·5%) in the pooled rabies control groups (event rate 0·21/100 child days) (table 1). In total, 72 of 338 participants (21·3%) had at least one episode of clinical malaria according to the primary case definition however, of these, 10 also had more than one episode. In a secondary analysis, VE against all clinical malaria episodes up to 6 months (as per the primary case definition), analysed using Cox regression models, was 56% (95% CI 24-74%; p=0·0035) for the delayed and 40% (95% CI 1-64%; p=0·045) for the monthly RH5.1/Matrix-M regimen (appendix pp 34-35).

As a secondary objective, we also analysed time to first episode of clinical malaria (as per the primary case definition) within 14 days to 3 months post-third vaccination using a Cox regression model. Here, a VE of 56% (95% CI 21-76%; p=0·0062) in the delayed regimen and 52% (95% CI 13-73%; p=0·015) in the monthly regimen was observed when comparing with the pooled control groups (appendix pp 44- 45). Here, 65 of 338 participants (19·2%) had at least one episode of malaria according to the primary case definition during this period.

We also analysed the time to first episode of clinical malaria, using a Cox regression model, according to secondary case definitions of clinical malaria during the 6-month period with i) a parasitaemia >0 per µL; here VE of 44% [95% CI 8-66%; p=0·023] was observed in the delayed group and 8% [95% CI -44-41%; p=0·73] in the monthly group; and ii) a parasitaemia >20,000 per µL; here VE of 69% [95% CI 35-85%; p=0·0017] was observed in the delayed group and 47% [95% CI 3-71%; p= 0·041] in the monthly group (table 1, figure 2B,C). In light of these results, additional post-hoc analyses were performed for case definitions of clinical malaria with i) parasitaemia >50,000 per µL; here VE of 81% [95% CI 44-94%; p=0·0026] was observed in the delayed group and 90% [95% CI 58-98%; p=0·0017] in the monthly group; and ii) parasitaemia >100,000 per µL; here VE of 83% [95% CI 23-96%; p=0·021] was observed in the delayed group and 91% [95% CI 33-99%; p=0·019] in the monthly group (table 1, figure 2D,E). Secondary analyses of all efficacy endpoints for the intention-to-treat population showed similar results to those for the per-protocol population (appendix pp 38-43, 46).

There were no SAEs, AEs of special interest (AESIs) or suspected unexpected serious adverse reactions (SUSARs) reported out to 6 months post-third vaccination. There were no safety concerns raised by the DSMB following review of 7 days’ data after vaccination of the first 100 participants, and no further safety reviews were required. Swelling was the most common local solicited AE, reported after 24/696 (3·4%) RH5.1/Matrix-M vaccinations, with significantly more swelling reported in the delayed RH5.1/Matrix-M group compared to the combined control groups (odds ratio [OR] 11·2, 95% CI 2·6- 49·4, p=0·0014). The most common systemic solicited AE was fever, reported after 97/696 (13·9%) RH5.1/Matrix-M vaccinations and 5/349 (1·4%) rabies vaccinations (table 2). There were significantly more fevers in both the delayed (OR 14·1, 95% CI 5·3-37·1, p<0·0001) and monthly (OR 9·7, 95% CI 3·7-25·9, p<0·0001) RH5.1/Matrix-M vaccination groups as compared to the control groups. No participants experienced febrile convulsions. The majority of solicited AEs were mild to moderate in severity. Four participants (1·7%) were reported to have severe pain following RH5.1/Matrix-M vaccination, compared to four (3·3%) following rabies vaccination. Three participants (1·3%) were reported to have severe fever and one participant (0·4%) reported to have severe loss of appetite following RH5.1/Matrix-M vaccination (appendix pp 47-48).

**Table 2:**
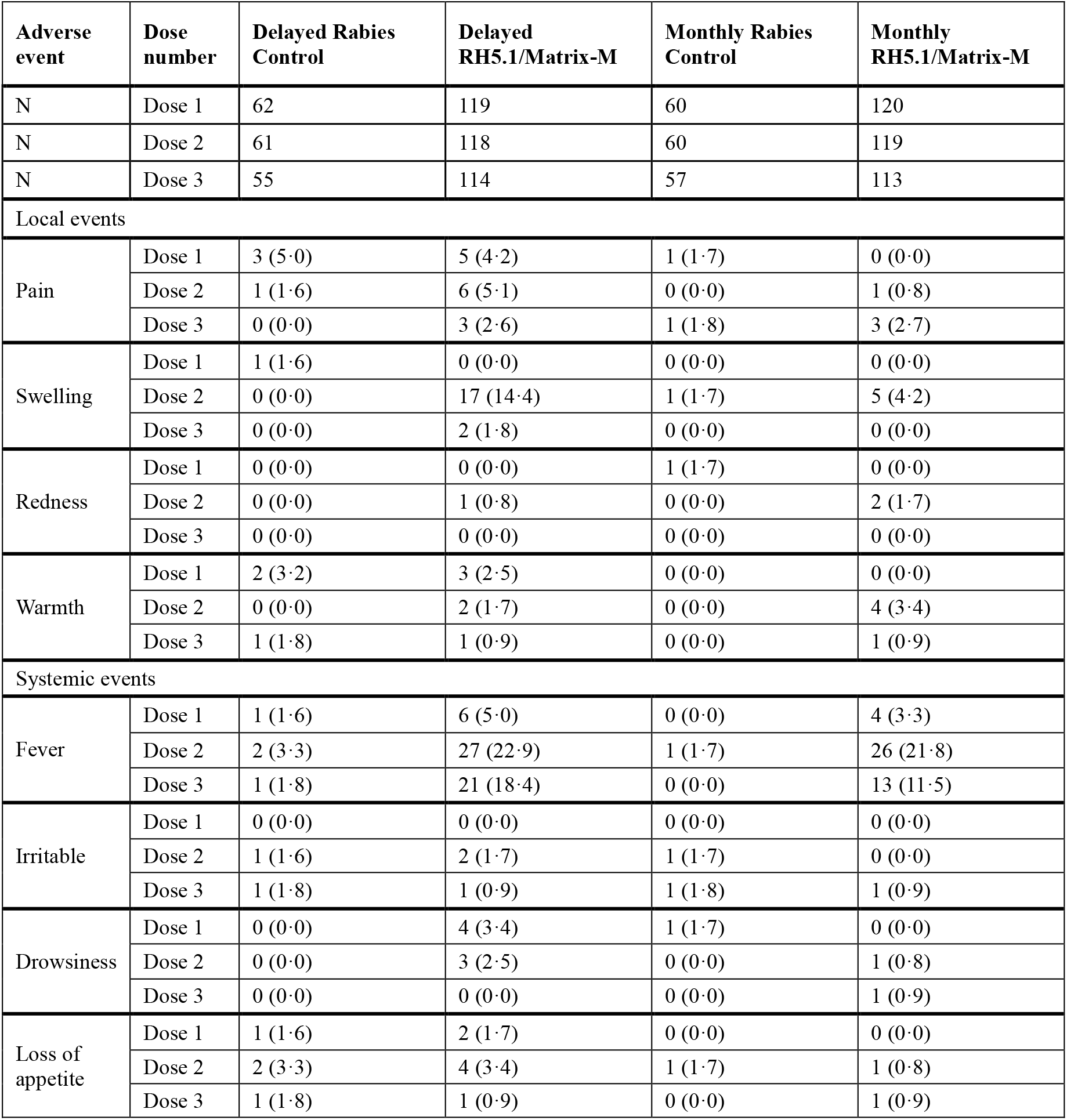
Solicited AEs within 7 days of vaccine dose across all groups.

| Adverse event | Dose number | Delayed Rabies Control | Delayed RH5.1/Matrix-M | Monthly Rabies Control | Monthly RH5.1/Matrix-M |
| --- | --- | --- | --- | --- | --- |
| N | Dose 1 | 62 | 119 | 60 | 120 |
| N | Dose 2 | 61 | 118 | 60 | 119 |
| N | Dose 3 | 55 | 114 | 57 | 113 |
| Local events |  |  |  |  |  |
| Pain | Dose 1 | 3 (5·0) | 5 (4·2) | 1 (1·7) | 0 (0·0) |
|  | Dose 2 | 1 (1·6) | 6 (5·1) | 0 (0·0) | 1 (0·8) |
|  | Dose 3 | 0 (0·0) | 3 (2·6) | 1 (1·8) | 3 (2·7) |
| Swelling | Dose 1 | 1 (1·6) | 0 (0·0) | 0 (0·0) | 0 (0·0) |
|  | Dose 2 | 0 (0·0) | 17 (14·4) | 1 (1·7) | 5 (4·2) |
|  | Dose 3 | 0 (0·0) | 2 (1·8) | 0 (0·0) | 0 (0·0) |
| Redness | Dose 1 | 0 (0·0) | 0 (0·0) | 1 (1·7) | 0 (0·0) |
|  | Dose 2 | 0 (0·0) | 1 (0·8) | 0 (0·0) | 2 (1·7) |
|  | Dose 3 | 0 (0·0) | 0 (0·0) | 0 (0·0) | 0 (0·0) |
| Warmth | Dose 1 | 2 (3·2) | 3 (2·5) | 0 (0·0) | 0 (0·0) |
|  | Dose 2 | 0 (0·0) | 2 (1·7) | 0 (0·0) | 4 (3·4) |
|  | Dose 3 | 1 (1·8) | 1 (0·9) | 0 (0·0) | 1 (0·9) |
| Systemic events |  |  |  |  |  |
| Fever | Dose 1 | 1 (1·6) | 6 (5·0) | 0 (0·0) | 4 (3·3) |
|  | Dose 2 | 2 (3·3) | 27 (22·9) | 1 (1·7) | 26 (21·8) |
|  | Dose 3 | 1 (1·8) | 21 (18·4) | 0 (0·0) | 13 (11·5) |
| Irritable | Dose 1 | 0 (0·0) | 0 (0·0) | 0 (0·0) | 0 (0·0) |
|  | Dose 2 | 1 (1·6) | 2 (1·7) | 1 (1·7) | 0 (0·0) |
|  | Dose 3 | 1 (1·8) | 1 (0·9) | 1 (1·8) | 1 (0·9) |
| Drowsiness | Dose 1 | 0 (0·0) | 4 (3·4) | 1 (1·7) | 0 (0·0) |
|  | Dose 2 | 0 (0·0) | 3 (2·5) | 0 (0·0) | 1 (0·8) |
|  | Dose 3 | 0 (0·0) | 0 (0·0) | 0 (0·0) | 1 (0·9) |
| Loss of appetite | Dose 1 | 1 (1·6) | 2 (1·7) | 0 (0·0) | 0 (0·0) |
|  | Dose 2 | 2 (3·3) | 4 (3·4) | 1 (1·7) | 1 (0·8) |
|  | Dose 3 | 1 (1·8) | 1 (0·9) | 0 (0·0) | 1 (0·9) |

For unsolicited AEs, 39 MedDRA terms were assigned in the 28 days following each of the three vaccinations and there were no significant differences in the number of events per group (appendix pp 49- 52). All were classified as unrelated to the vaccines, with the exception of a single episode of moderate fever in the delayed third-dose group within 28 days of vaccination. There were no cases of severe anaemia (haemoglobin <5·0g/L), no participants requiring blood transfusion, and no significant differences in the frequency of moderate anaemia (haemoglobin <8·0g/L) between the groups (appendix 53). There were no cases of severe malaria in any participant in the 6 months post-third vaccination.

At baseline, almost all participants showed background level anti-RH5.1 serum IgG antibody responses. At 14 days post-third vaccination, responses remained comparable to baseline in the combined rabies vaccine control groups. In contrast, high responses were seen in the delayed regimen RH5.1/Matrix-M group (geometric mean anti-RH5.1 IgG concentration 837µg/mL; inter-quartile range [IQR] 326-2200); these were significantly higher as compared to the monthly regimen group (geometric mean 626µg/mL; IQR 222-1455; p=0·0006) (figure 3A). *In vitro* functional anti-parasitic activity was assessed by the GIA assay at a total purified IgG concentration of 2·5 mg/mL at 14 days post-third vaccination. Samples from control participants showed negligible GIA (<20%), apart from three participants who had GIA of >50%. Mean GIA in the delayed regimen RH5.1/Matrix-M group was 79·0% (standard deviation [SD] 14·3), significantly higher than the mean GIA in the monthly group of 74·2% (SD 15·9; p=0·016) (figure 3B, appendix pp 54-55).

**Figure 3:**
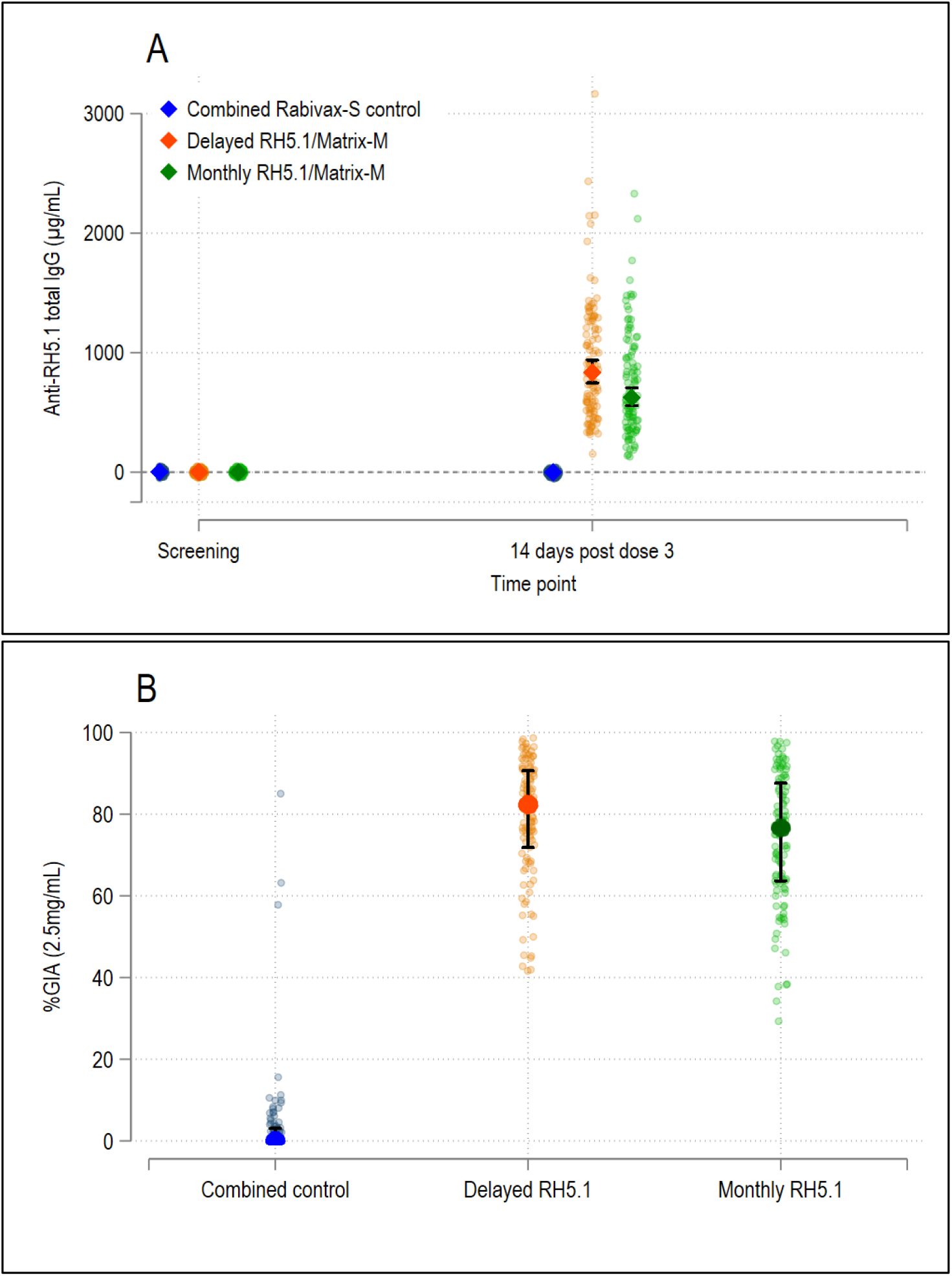
RH5.1/Matrix-M humoral immunogenicity.

## Discussion

We report that the standalone blood-stage vaccine candidate RH5.1/Matrix-M, delivered in a delayed third dose (0-1-5 month) regimen, shows significant efficacy of 55% (95% CI 20-75%) against clinical malaria in the target population of African children over 6 months’ follow-up. The vaccine was also well-tolerated and no SAEs were reported out to 6-months following the third vaccination. Data now reported for a total of 109 adults, 305 children and 18 infants vaccinated with RH5-based vaccines in the UK, Tanzania and Burkina Faso^14,16–18^ all show similar safety and tolerability profiles, whilst Matrix-M adjuvant is now licensed in vaccines for malaria and Covid-19.^3,21^ On-going phase 1/2 trials continue to monitor the safety of RH5.1/Matrix-M vaccination.

Here we studied RH5.1/Matrix-M efficacy for the first time in an area of seasonal malaria transmission, with vaccinations completing part-way through the malaria season. Previously published field efficacy trials of *P. falciparum* blood-stage vaccine candidates, targeting a variety of antigens including AMA1, MSP1, MSP2, GLURP, MSP3 and RESA, all yielded disappointing results, reporting either no or minimal clinical efficacy, or in some cases evidence of strain-specific efficacy linked to target antigen polymorphism.^5–9^ Given these studies were performed over the last 25 years in different settings with different transmission patterns and in children of different age ranges, it is not possible to directly compare with our study. However, our results demonstrate that a standalone blood-stage vaccine can achieve significant efficacy against clinical malaria in 5-17 month old children, in line with the approved age range for use of the pre-erythrocytic vaccines RTS,S/AS01 and R21/Matrix-M, and now enabling future assessment of combination malaria vaccine strategies targeting two stages of the parasite’s lifecycle. Indeed, a second-generation multi-stage paediatric vaccination strategy offers hope for higher and more durable efficacy against clinical malaria, especially if the pre-erythrocytic and blood-stage components act additively, or even synergistically.

SMC was given to children in the study area as part of a programme by local health services, as per national policy recommendation, but coverage was suboptimal. Our study documented SMC uptake, but did not deliver any further SMC. It will be important in the future to test RH5.1/Matrix-M efficacy in settings where there is no SMC, and to also determine in seasonal settings whether combination of blood-stage and pre-erythrocytic vaccines can achieve a better outcome (than a pre-erythrocytic vaccine alone) as indicated by studies of RTS,S/AS01 and SMC.^22^

Our study was not powered to show a difference in VE between the 0-1-2 and 0-1-5-month delivery regimens with RH5.1/Matrix-M. However, the VE for the 0-1-5 month regimen was higher than for the 0-1-2 month regimen, and this is consistent with the 0-1-5-month regimen inducing higher IgG concentrations and GIA. The difference in VE was larger at 6 months than 3 months, suggesting the 0-1- 5-month regimen may offer more durable protection. Previous trials of RTS,S/AS01 reported improved efficacy against malaria challenge in healthy US adults when using a 0-1-7-month regimen with antigen and adjuvant fractionated for the delayed third dose (as opposed to 0-1-2-month dosing),^23^ however, this did not translate to improved field efficacy in 5-17 month old children.^24^. In line with these observations, our previous phase 1b trial data with RH5.1/Matrix-M in Tanzanian children suggested that a delayed (full) third dose, as opposed to monthly dosing or a delayed fractional third dose, may induce more robust and durable antibody responses.^18^ On-going analyses in this trial will thus continue to investigate whether the delayed third dose regimen induces more durable immunity in contrast to monthly dosing.

Notably, our secondary and post-hoc analyses showed lower VE (as compared to the primary endpoint) when we used the secondary clinical malaria case definition of any parasitaemia (>0 parasites/µL) but increasingly improved efficacy when we used secondary or additional malaria case definitions with a higher parasitaemia cut-off of >20,000, >50,000 or >100,000 parasites/µL. The delayed third dose RH5.1/Matrix-M regimen showed significant efficacy against all definitions, whilst the monthly regimen only reached significance at the higher cut-offs consistent with the more modest performance of this regimen but nonetheless suggesting biological effect. These data appear in line with research in animal vaccination and challenge models of malaria,^13^ whereby blood-stage malaria vaccines can reduce peak parasitaemia. This would also not be expected to occur with pre-erythrocytic vaccines; indeed, post-hoc analysis of data from the phase 3 trial of R21/Matrix-M^3^ showed almost identical VE for all the cut-off levels of parasitaemia analysed here (Adrian Hill, personal communication). Consequently, our data show that RH5.1/Matrix-M can partially protect against clinical malaria but can also reduce blood-stage parasitaemia in clinical cases. This may have implications for prevention of severe or life-threatening disease in the real-world setting when pre-erythrocytic- and/or blood-stage vaccine-induced immunity to clinical malaria fails or wanes.

Solicited AE rates observed with RH5.1/Matrix-M compare favourably to those seen in the phase 3 trials of R21/Matrix-M and RTS,S/AS01.^2,3^ Swelling at the injection site, the most common local solicited AE in this trial, occurred following 4% of vaccinations with R21/Matrix-M and 10% with RTS,S/AS01, in comparison to 3% with RH5.1/Matrix-M. For R21/Matrix-M and RTS,S/AS01, the most common local solicited AE was pain, occurring following 19% and 12% of vaccinations respectively, in comparison to 3% with RH5.1/Matrix-M. Fever was the most common systemic solicited AE with all vaccines, occurring following 47% of vaccinations with R21/Matrix-M and 31% with RTS,S/AS01, in comparison to 14% with RH5.1/Matrix-M.

The absence of RH5 serum antibody responses at baseline or in the control groups at 14 days post-third vaccination is consistent with the known sero-epidemiology and sequence conservation of RH5, suggesting this antigen is not a dominant target of naturally-acquired malaria immunity.^10,16,25^ In contrast, the RH5.1/Matrix-M vaccine candidate was highly immunogenic for functional anti-RH5.1 serum IgG antibody across both dosing regimens, with the delayed third dose regimen showing small but significant improvements in the ELISA and GIA responses (as compared to the monthly regimen) 14 days post-third vaccination, in line with our data seen in Tanzanian children in the RH5.1/Matrix-M phase 1b study.^18^ The mean GIA observed in both RH5.1/Matrix-M groups, and over 80% of individual children, at this timepoint also exceeded 60% GIA measured at 2·5 mg/mL total IgG (appendix p 54-55) – a threshold level we previously reported as required for protection following RH5 vaccination and *P. falciparum* blood-stage challenge in *Aotus* monkeys.^13,15^ These data are thus consistent with both RH5.1/Matrix-M regimens showing significant efficacy against clinical malaria in the first 3 months post-third vaccination. Analysis of the kinetic of both groups’ immune responses beyond this peak post-vaccination timepoint are ongoing, but the efficacy data after 3 months suggest that differences may be seen with respect to serum antibody durability and/or the possibility for natural boosting of the vaccine-induced response.

This study has limitations, including the completion of vaccine doses part-way through the malaria season. It is possible that VE might be different if administered earlier, i.e. with the primary vaccination series (all three doses) being completed prior to the season. Another limitation that comes from administering the vaccine part-way through the malaria season is that there was insufficient follow up time to observe many children having multiple episodes. It is possible that naturally-acquired immunity may interact with vaccine-induced immunity, protecting children from subsequent episodes, but the current analysis was unable to investigate this. Nonetheless, follow-up of the VAC091 trial is continuing to determine efficacy at 12 months post-third vaccination, and to assess the durability of the vaccine-induced immune response and the potential impact of natural malaria exposure. We will also administer a fourth (booster) dose of vaccine at 12 months to Groups 1-4 to enable efficacy monitoring over a second year of follow-up. We also limited the age range of participants in this trial to 5-17 months, to align with earlier studies of RTS,S/AS01 and R21/Matrix-M. A wider age range, inclusive of younger infants and older children, will be covered in future trials. We have not yet assessed RH5.1/Matrix-M delivered in an age-based (non-seasonal) administration schedule, or in sites with lower or higher levels of perennial malaria transmission as compared to the seasonal setting in Nanoro; this will be addressed in future studies. We have also not yet formally analysed our immunological datasets for correlates of protection; this, along with assessment for any evidence of *P. falciparum* strain-selection in vaccinees versus controls, remains the focus of ongoing work.

## Contributors

JS, SES, SJD and AMM conceived the trial. HT was the trial principal investigator, AMM the chief investigator, and SJD the senior laboratory investigator. HMN, JS, AS, SD, OT, TR, FO, EO, KCSD, NAK, ZMJC, MK, MAB, FK, HS, CMN, DP, AD, CAL, KM, SES, HT and AMM contributed to the implementation of the study and data collection. SS and JB analysed data and completed statistical analysis. HMN, SD, JSC, AML, KS and RR performed project management. LDWK, US and KS assisted with vaccine provision. HMN, JS, KM, SES, SJD, HT and AMM interpreted data, contributed to writing the manuscript and have accessed and verified the data.

## Data sharing

Data associated with this study are present in the paper or appendix and will be available after the end of the study upon reasonable request that should be directed to the corresponding author. Proposals will be reviewed and approved by the Sponsor, Chief Investigator, and collaborators. After approval of a proposal, data can be shared through a secure online platform after signing a data access agreement. Any shared data will be de-identified.

## Acknowledgments

We thank all the trial participants and their parents, and the independent DSMB for overseeing the trial. This work was funded in part by the European and Developing Countries Clinical Trials Partnership (EDCTP) Multi-Stage Malaria Vaccine Consortium (MMVC) [RIA2016V-1649-MMVC]; a Wellcome Trust Translation Award [205981/Z/17/Z], the UK Medical Research Council [MR/K025554/1 and MR/V038427/1, the latter UK-funded award is carried out in the frame of the Global Health EDCTP3 Joint Undertaking]; the National Institute for Health and Care Research (NIHR) Oxford Biomedical Research Centre (BRC), the views expressed are those of the authors and not necessarily those of the NHS, the NIHR, or the Department of Health; Matrix-M adjuvant and rabies vaccine was supplied by SIIPL; the GIA work was supported by the Division of Intramural Research, National Institute of Allergy and Infectious Diseases (NIAID) and by an Interagency Agreement [AID-GH-T-15-00001] between the United States Agency for International Development (USAID) Malaria Vaccine Development Program (MVDP) and NIAID. The findings and conclusions are those of the authors and do not necessarily represent the official position of USAID. CMN held a Wellcome Trust Sir Henry Wellcome Postdoctoral Fellowship [209200/Z/17/Z]. SJD is a Jenner Investigator. All authors had full access to all the data in the study and had final responsibility for the decision to submit for publication.

## Figure Legends

Figure Legends Figure 1: Trial profile.

Participants allocated to the delayed regimen received vaccinations at 0-1-5 months. Participants allocated to the monthly regimen received vaccinations at 0-1-2 months. Participants were aged 5-17 months at enrolment (first day of vaccination). MM = Matrix-M.

Table 1: Analysis of time to first episode of clinical malaria from 14 days to 6 months post-third vaccination.

Cox regression models were used to analyse the primary endpoint of time to first episode of clinical malaria based on the per-protocol population at the 6-month analysis timepoint. Clinical malaria is defined as measured axillary temperature ≥37·5°C and/or reported history of fever within the last 24 hours, and *P. falciparum* asexual parasitaemia concentration according to each case definition. Vaccine Efficacy = 1 – hazard ratio. 95% CI = 95% Confidence Interval. * Secondary analysis adjusted vaccine efficacy: includes covariates of total number of rounds of SMC received, ITN use (adequate or not) the night before the screening visit, and age categories 5-8 months, 9-12 months, 13-17 months. Excludes 5 children with missing data on covariates: 1 in the Delayed Rabies arm, 3 in the Delayed RH5.1/Matrix-M arm and 1 in the Monthly RH5.1/Matrix-M arm. ** Event rate per 100 days (where event is the first episode of clinical malaria) is also reported for information only, calculated as: (the number of events / [child days/100]).

Figure 2: Kaplan-Meier curves showing time to first episode of clinical malaria from 14 days to 6 months post-third vaccination.

The primary analysis was based on the per-protocol population. (**A**) Primary case definition of clinical malaria with parasitaemia >5000 parasites/µL. (**B**) Secondary case definition of clinical malaria with parasitaemia >0 parasites/µL, or (**C**) >20,000 parasites/µL. (**D**) Additional case definition of clinical malaria with parasitaemia >50,000 parasites/µL, or (**E**) >100,000 parasites/µL.

Table 2: Solicited AEs within 7 days of vaccine dose across all groups.

By number of doses (n and (%)). Includes all children in the intention-to-treat sample who received at least one dose of vaccine. N = number of participants who received each dose.

Figure 3: RH5.1/Matrix-M humoral immunogenicity.

Immunological outcomes in study participants in the per-protocol sample. (**A**) Anti-RH5.1 serum IgG responses by vaccination group at baseline (screening) and day 14 post-vaccine dose three. Individual anti-RH5.1 total IgG antibody concentrations (dots) and geometric mean with 95% CIs (diamonds with black bars). N=111 children for the Rabies (Rabivax-S) delayed and monthly groups combined, N=113 for the Delayed RH5.1/Matrix-M group, and N=107 for the Monthly RH5.1/Matrix-M group. (**B**) Percentage *in vitro* GIA of 3D7 clone *P. falciparum* parasites by vaccination group, using 2·5mg/mL total IgG purified from serum taken on day 14 post-vaccine dose three. Individual percentage inhibition figures (small dots) and median and interquartile range (large dots and black bars). N=97 children for the Rabies (Rabivax-S) delayed and monthly groups combined, N=113 children for the Delayed RH5.1/Matrix-M group, and N=108 for the Monthly RH5.1/Matrix-M group.

