## appendix for "Safety and efficacy of the blood-stage malaria vaccine RH5.1/Matrix-M in Burkina Faso: interim results of a double-blind, randomised, controlled phase 2b trial in children"

### Supplementary Appendix

### 1. Contributions to the VAC091 Trial

#### 1.1 Acknowledgments

We thank all the trial participants and their parents, the Nanoro Health District authorities, the CMA Saint Camille de Nanoro hospital, and all the research staff at the Unité de Recherche Clinique de Nanoro. The authors are grateful to Rebecca Ashfield, Sophie Barnett, Jordan Barrett, Jennifer Bryant, Mehreen Datoo, Hannah Davies, Adrian Hill, Jing Jin, David Pulido, Jack Quaddy, Subarna Sriskantharajah, Lana Strmecki, Daniela Tomescu, Cheryl Turner and Hilary Vitkus for assistance and Caroline Hosking for arranging contracts (University of Oxford); Catherine Green, Abdou Tahiri-Alaoui, Emma Bolam, Richard Tarrant and Tanja Brenner (Clinical Biomanufacturing Facility, University of Oxford); Jenny Reimer and Cecilia Carnrot (Novavax); and the independent DSMB for their insightful safety oversight (Brian Angus, Karim Manji, Roma Chilengi, Kwaku Poke Asante, William Macharia and Greg Fegan).

### 2 Methods

#### 2.1 Study Design

VAC091 is a phase 2b double-blinded, block-randomised, controlled trial. It is being conducted according to the principles of the current revision of the Declaration of Helsinki 2013 and in full conformity with the ICH guidelines for Good Clinical Practice (GCP). It is sponsored by the University of Oxford and was approved by the Oxford Tropical Research Ethics Committee in the UK (reference 3-23) and the National Ethical Committee of the Ministry of Health (reference 2022-12-256) and Regulatory Authority (reference 20230208/MSHP/SG/ANRP/DHEC/MIK) in Burkina Faso, and was registered on ClinicalTrials.gov (NCT05790889). The trial is being conducted in Burkina Faso by the Institut de Recherche en Sciences de la Santé – Unité de Recherche Clinique de Nanoro/Clinical Research Unit of Nanoro (IRSS-URCN/CRUN) at the Siglé trial site located within the Nanoro Health District catchment area.

We report here the safety, immunogenicity and efficacy of vaccination with RH5.1 soluble protein given with 50 µg Matrix-M adjuvant[1,2] in monthly and delayed third dose regimens up until 6 months post-third study vaccination. The Consolidated Standards of Reporting Trials (CONSORT) guidelines were followed.

#### 2.2 Study Site

IRSS-URCN is situated approximately 90 km north-west of Ouagadougou, the capital city of Burkina Faso. The VAC091 study participants were recruited at the Siglé trial site located within the Nanoro Health District catchment area. The health district covers 24 villages where health care is provided by 11 peripheral health posts and one referral hospital. The population of the district is just over 63,000 inhabitants.

In Burkina Faso, malaria is endemic; transmission occurs throughout the year, with a peak during the rainy season (June to November). *Plasmodium falciparum* is responsible for more than 90 % of all clinical malaria cases and the major mosquito vectors are *Anopheles gambiae, An. arabiensis and An. funestus*. Children under five years and pregnant women are the populations at highest risk; during the six months when transmission reaches a peak, individuals in these age-brackets may suffer multiple malaria episodes. There were almost 12 million recorded cases and over 4,200 deaths attributed to malaria in 2022.[3] From recent surveys, approximately 64 % of the population had access to an insecticide-treated bednet (ITN).[3] Seasonal malaria chemoprevention (SMC) in children aged from 3 to 59 months was implemented country-wide in 2017 (http://cns.bf/IMG/pdf/annuaire_ms_2017.pdf) with pilot implementation in some districts from 2014. However, there is no implementation of indoor residual spraying (IRS) with insecticide in the area.

#### 2.3 Sample Size

The sample size was calculated with the objective of detecting a vaccine efficacy of 50% compared to the pooled control groups (i.e., a hazard ratio of 0.5). The VAC091 study was designed with conservative estimates of malaria disease rates to deliver reliably, and with high power, the primary endpoint of efficacy against clinical malaria disease over six months for each of the vaccination regimens. Using data from the R21/Matrix-M phase 3 study (VAC078; ClinicalTrials.gov NCT04704830),[4] which was conducted at the same site, in the control arm there were 1·2 events per child in the first 6 months of follow up which meant that 104 children per arm were required for 90% power to detect a difference between arms. Loss to follow up in that study was low, at 4%, therefore 108 children per arm would need to be recruited. The group sizes in VAC091 of 120 therefore still gave over 80% power even if the rate of malaria decreased to 0·8 events per child in the control groups.

#### 2.4 Participants and Recruitment

Healthy children aged 5-17 months residing in Siglé, Nanoro District, Burkina Faso were eligible for inclusion in the VAC091 study. The guardian of each participant signed or thumb-printed an informed consent form at the screening visit and consent was verified before each vaccination. Enrolment was into four groups (**Table S1**). The delayed third dose regimen groups were recruited as an initial cohort and the monthly third dose regimen groups were recruited as a second cohort, with the final vaccination for all groups given contemporaneously.

There was a planned Data Safety Monitoring Board (DSMB) review after recruitment of the first 100 participants in the delayed third dose cohort. This was a review of seven days of safety data with no trial pause. There were no other pre-specified DSMB review points.

##### 2.4.1 Inclusion and Exclusion Criteria

###### *2.4.1.1 Inclusion Criteria*

Only participants who met all the inclusion criteria were enrolled into the VAC091 trial:

- Healthy child aged 5-17 months at the time of first study vaccination;
- Parent/guardian provides signed/thumb-printed informed consent;
- Child and parent/guardian resident in the study area villages and anticipated to be available for vaccination and follow-up for 12 months following last dose of vaccination.

###### *2.4.1.2 Exclusion Criteria*

Participants were excluded if ANY of the following applied:

- Clinically significant congenital abnormalities as judged by the Principal Investigator (PI) or other delegated individual;
- Clinically significant skin disorder (psoriasis, contact dermatitis etc.), cardiovascular disease, respiratory disease, endocrine disorder, liver disease, renal disease, gastrointestinal disease, neurological illness as judged by the PI or other delegated individual;
- Weight-for-age Z score of less than -3 or other clinical signs of malnutrition;
- History of allergic reaction, significant IgE-mediated event, or anaphylaxis to immunisation;
- History of allergic disease or reactions likely to be exacerbated by any component of the vaccines;
- Sickle cell disease;

- Clinically significant laboratory abnormality as judged by the study clinician;
- Administration of immunoglobulins and/or any blood products within the three months preceding the planned administration of the vaccine candidate;
- Receipt of any vaccine in the 14 days preceding enrolment, or planned receipt of any other vaccine within 28 days following each study vaccination;
- History of vaccination with another malaria vaccine;
- Participation in another research study involving receipt of an investigational product in the 30 days preceding enrolment, or planned use during the study period;
- Known maternal HIV infection;
- Any confirmed or suspected immunosuppressive or immunodeficient state, including HIV infection; asplenia; recurrent, severe infections and chronic (more than 14 days) immunosuppressant medication within the past 6 months (for corticosteroids, this will mean prednisone, or equivalent, ≥ 0·5 mg/kg/day; inhaled and topical steroids are allowed);
- Any significant disease, disorder or situation which, in the opinion of the Investigator, may either put the participants at risk because of participation in the trial, or may influence the result of the trial, or the participant's ability to participate in the trial.

##### 2.5 Randomisation and Blinding

Eligible participants whose carers had consented to participation and had completed the baseline assessment were randomised. Children were stratified by age and sex and randomised to Groups 1 and 2 at an allocation ratio of 1:2, or to Groups 3 and 4 by the same ratio. Allocation was carried out using sequentially numbered opaque sealed envelopes. These were prepared and sealed by an independent statistician using a random allocation list using block randomisation with variable block sizes. After participant eligibility was confirmed by two Investigators, the unblinded study pharmacist was allowed to open an envelope.

The unblinded pharmacists did not participate in any of the study endpoint data evaluation. All other site personnel, the central study team and the laboratory team involved in the immunogenicity analysis were blinded to the study vaccine administration.

The Local Safety Monitor (LSM) and DSMB, who are independent from the study team, were also provided with the randomisation sequence so that, if deemed necessary for reasons such as safety, the LSM or DSMB would be able to unblind the specific enrolled participant without revealing the study group to the Investigators.

#### 2.6 Safety Analysis

Following each vaccination, each participant was visited at home on days 1 to 6 by a community health worker for assessment and any solicited and unsolicited adverse events (AEs) were recorded. At days 14 and 28 post-vaccination, participants were seen at the clinical research facility. In addition, participants attended the facility for follow-up at 2 and 6 months post-third vaccination. At any other times during the study period, participants were visited at home once a month. The full follow-up schedule is shown in **Table S4** and **Table S5**.

Solicited AEs were collected for 7 days post-vaccination (day of vaccination and 6 subsequent days) and unsolicited AEs were collected for 28 days post-vaccination (day of vaccination and 27 subsequent days). Serious adverse events (SAEs) were collected throughout the study period. Safety blood samples (full blood count, bilirubin, alanine aminotransferase [ALT] and creatinine) were taken at screening, 14 days post-second vaccination, day of third vaccination (delayed third dose regimen only) and six months post-third vaccination. Temperature was taken at all visits apart from the screening visit.

##### 2.6.1 Adverse Event Causality and Grading

Solicited AEs were graded by Investigators according to the scale in **Table S2** and unsolicited AEs were graded according to **Table S3**. Abnormal laboratory results were assessed for clinical significance and any clinically significant abnormalities graded according to the Division of AIDS grading scale.[5]

Solicited AEs were defined as being at least possibly related to vaccination. The causal relationship between unsolicited AEs and SAEs and the product was evaluated by the PI or PI-delegated clinician. This interpretation was based on the type of event, the relationship of the event to the time of vaccine administration, and the known biology of the vaccines. The following guidelines were used for assessing the causal relationship:

- No relationship:
  - No temporal relationship to study product; *and*
  - Alternate aetiology (clinical state, environmental or other interventions); *and*
  - Does not follow known pattern of response to study product
- Unlikely relationship:
  - Unlikely temporal relationship to study product; *and*
  - Alternate aetiology likely (clinical state, environmental or other interventions); *and*
  - Does not follow known typical or plausible pattern of response to study product
- Possible relationship:
  - Reasonable temporal relationship to study product; *or*
  - Event not readily produced by clinical state, environmental or other interventions; *or*
  - Similar pattern of response to that seen with other vaccines
- Probable relationship:
  - Reasonable temporal relationship to study product; *and*
  - Event not readily produced by clinical state, environment, or other interventions; *or*
  - Known pattern of response seen with other vaccines
- Definite relationship:
  - Reasonable temporal relationship to study product; *and*
  - Event not readily produced by clinical state, environment, or other interventions; *and*

- Known pattern of response seen with other vaccines

Unsolicited AEs were classified according to MedDRA (version 27·0).

#### 2.7 Study Objectives

All study objectives in the VAC091 protocol are included below, however, not all are reported in this paper. This paper reports interim results including the primary objective efficacy data, and available safety data, for Groups 1 to 4 out to 6 months post-third vaccination (17th March 2024). The remaining endpoints will be reported elsewhere following the end of the trial once these data are available (expected in December 2025). These include endpoints relating to protocol amendments that have since incorporated a fourth (booster) dose of RH5.1/Matrix-M, without or with a 3-dose series of the approved R21/Matrix-M vaccine, at 12 months post-third RH5.1/Matrix-M vaccination in Groups 2 and 4, respectively; along with additional safety and immunogenicity cohorts (new Groups 5 and 6) vaccinated with a candidate RH5-based virus-like particle vaccine (RH5.2-VLP/Matrix-M).[6]

##### 2.7.1 Primary Objectives

###### *2.7.1.1 Efficacy against clinical malaria (Groups 1-4)*

- To assess the protective efficacy against clinical malaria of RH5.1 with Matrix-M in 5-17 month old children living in a malaria-endemic area, for 6 months after the third vaccination (reported here).

###### *2.7.1.2 Safety*

- To assess the safety and reactogenicity of RH5.1 with Matrix-M, RH5.2-VLP with Matrix-M and R21 with Matrix-M in 5-17 month old children living in a malaria-endemic area, in the month following each vaccination, and for 12 months after administration of the third dose of vaccine (Groups 1-6) and for 12 months after a fourth (booster) dose of RH5.1 with Matrix-M (Groups 1-4). Here we only report RH5.1/Matrix-M and rabies vaccine control groups (Groups 1-4) data out to 6 months post-third vaccination.

##### 2.7.2 Secondary Objectives

###### 2.7.2.1 *Immunogenicity*

- To assess the humoral immunogenicity of RH5.1 with Matrix-M and RH5.2-VLP with Matrix-M in 5-17 month old children living in a malaria-endemic area (only RH5.1/Matrix-M data reported here).
- To assess for any immune interference between R21 and RH5.1 with Matrix-M in 5-17 month old children living in a malaria-endemic area (Groups 3-4) (not reported here).

###### 2.7.2.2 *Efficacy against clinical malaria (Groups 1-4)*

- To assess the protective efficacy against clinical malaria of RH5.1 with Matrix-M in 5-17 month old children living in a malaria-endemic area, for 3 months and 12 months after the third vaccination (only 3-month data reported here).
- To assess the protective efficacy against clinical malaria of RH5.1 with Matrix-M plus/minus R21 with Matrix-M in 5-17 month old children living in a malaria-endemic area, for 3, 6 and 12 months after the fourth (booster) dose of RH5.1 in Matrix-M (not reported here).

###### 2.7.2.3 *Efficacy against asymptomatic infection (Groups 1-4)*

- To assess the protective efficacy against asymptomatic *P. falciparum* infection of RH5.1 with Matrix-M in 5-17 month old children living in a malaria-endemic area, by quantitative polymerase chain reaction (qPCR) at 2, 6 and 12 months after administration of the third dose of vaccine (not reported here).
- To assess the protective efficacy against asymptomatic *P. falciparum* infection of RH5.1 with Matrix-M plus/minus R21 with Matrix-M in 5-17 month old children living in a malaria-endemic area, by qPCR at 2, 6 and 12 months after administration of a fourth (booster) dose of RH5.1 with Matrix-M (not reported here).

###### *2.7.2.4 Efficacy against gametocytaemia (Groups 1-4)*

- To assess the protective efficacy against gametocytaemia of RH5.1 with Matrix-M in 5-17 month old children living in a malaria-endemic area, by quantitative reverse-transcriptase PCR (qRT-PCR) at 2, 6 and 12 months after administration of the third dose of vaccine (not reported here).
- To assess the protective efficacy against gametocytaemia of RH5.1 with Matrix-M plus/minus R21 with Matrix-M in 5-17 month old children living in a malaria-endemic area, by qRT-PCR, at 2, 6 and 12 months after administration of a fourth (booster) dose of RH5.1 with Matrix-M (not reported here).

###### *2.7.2.5 Efficacy against anaemia (Groups 1-4)*

- To assess the protective efficacy of RH5.1 with Matrix-M vaccination regimens against prevalent moderate or severe anaemia at 6 and 12 months after administration of the third dose of vaccine (only 6-month data reported here).
- To assess the protective efficacy of RH5.1 with Matrix-M plus/minus R21 with Matrix-M vaccination regimens against prevalent moderate/severe anaemia at 6 and 12 months after administration of a fourth (booster) dose of RH5.1 with Matrix-M (not reported here).

##### 2.7.3 Exploratory Objectives

###### *2.7.3.1 Efficacy against severe malaria (Groups 1-4)*

- To assess the protective efficacy against severe malaria of RH5.1 with Matrix-M in 5-17 months old children living in a malaria-endemic area, at 6 and 12 months after administration of the third dose of vaccine (only 6-month data reported here).
- To assess the protective efficacy against severe malaria of RH5.1 with Matrix-M plus/minus R21 with Matrix-M in 5-17 months old children living in a malaria-endemic area, at 6 and 12 months after administration of a fourth (booster) dose of RH5.1 with Matrix-M (not reported here).

###### *2.7.3.2 Efficacy of two doses against clinical malaria in a delayed third dose regimen (Groups 1-2)*

- To assess the protective efficacy against clinical malaria of RH5.1 with Matrix-M in 5-17 months old children living in a malaria-endemic area, between the second and third doses in a delayed third dose regimen (not reported here).

###### *2.7.3.3 Immunogenicity*

- To evaluate cellular immunogenicity and other exploratory immunological endpoints (not reported here).

###### *2.7.3.4 Efficacy against anaemia (Groups 1-4)*

- To assess the protective efficacy of RH5.1 with Matrix-M vaccination regimens against incident severe anaemia for 6 months after administration of the third dose of vaccine (reported here).
- To assess the protective efficacy of RH5.1 with Matrix-M plus/minus R21 with Matrix-M vaccination regimens against incident severe anaemia for 6 months after administration of a fourth (booster) dose of RH5.1 with Matrix-M (not reported here).
- To assess the protective efficacy of RH5.1 with Matrix-M vaccination regimens in preventing the need for blood transfusion for 6 months after administration of the third dose of vaccine (reported here).
- To assess the protective efficacy of RH5.1 with Matrix-M plus/minus R21 with Matrix-M vaccination regimens in preventing the need for blood transfusion for 6 months after administration of a fourth (booster) dose of RH5.1 with Matrix-M (not reported here).

#### 2.8 Endpoints

Endpoints were evaluated at the time-points indicated in **Table S4** and **Table S5**.

##### 2.8.1 Efficacy against clinical malaria

For Groups 1 and 2, assessment for efficacy outcomes will begin from 14 days after third and fourth study vaccinations (RH5.1/Matrix-M or rabies) until the specified timepoint. This excludes the exploratory objective of the protective efficacy between the second and third dose in the delayed regimen, which will be collected from 14 days after the second study vaccination until 13 days after the third study vaccination.

For Groups 3 and 4, assessment for efficacy outcomes will begin from 14 days after the third vaccination in year 1 (RH5.1/Matrix-M or rabies) and from 14 days after the seventh vaccination in year 2 (fourth dose of RH5.1/Matrix-M or rabies) until the specified timepoint.

*Primary case definition:*

- Presence of axillary temperature ≥37.5°C and/or history of fever in the last 24 hours AND *P. falciparum* parasite density >5000 asexual forms/µL.

*Secondary case definitions:*

- Presence of axillary temperature ≥37.5°C and/ or history of fever within the last 24 hours AND EITHER *P. falciparum* asexual parasitaemia >0 parasites/µL.
- Presence of axillary temperature ≥37.5°C and/ or history of fever within the last 24 hours AND *P. falciparum* asexual parasitaemia >20,000 parasites/µL.
- Comparison of mean parasite density in each study arm (not reported here).

After their third vaccination, participants were visited by field workers approximately every 30 days until 6 months after the third vaccination and a blood spot was taken for parasite quantification and genotyping. If they had a temperature of ≥37·5°C and/or history of fever within the last 24 hours, blood sampling was also performed for blood film microscopy to detect *Plasmodium* spp. This was done by experienced microscopists and all slides were double read. In the case of any discrepancy a third read was requested.

###### 2.8.2 Efficacy against asymptomatic infection

- The proportion of participants in each study arm that show presence of parasite density >5000 asexual forms/µL as measured by qPCR from 14 days to 2, 6 and 12 months after administration of the third and fourth doses of the RH5.1/Matrix-M or rabies control vaccine *PLUS* presence of axillary temperature <37.5°C and absence of history of fever within the last 24 hours.
- The proportion of participants in each study arm that show presence of parasite density >0 asexual forms/µL as measured by qPCR from 14 days to 2, 6 and 12 months after administration of the third and fourth doses of the RH5.1/Matrix-M or rabies control vaccine *PLUS* presence of axillary temperature <37.5°C and absence of history of fever within the last 24 hours.
- The parasite density in those with >0 asexual forms/µL as measured by qPCR from 14 days to 2, 6 and 12 months after administration of the third and fourth doses of the RH5.1/Matrix-M or rabies control vaccine *PLUS* presence of axillary temperature <37.5°C and absence of history of fever within the last 24 hours.

###### 2.8.3 Efficacy against gametocytaemia

- The proportion of participants in each study arm that show presence of gametocytes >0 gametocytes/µL as measured by qRT-PCR from 14 days to 2, 6 and 12 months after administration of the third and fourth doses of the RH5.1/Matrix-M or rabies control vaccine *PLUS* presence of axillary temperature <37.5°C and absence of history of fever within the last 24 hours.

###### 2.8.4 Efficacy against severe malaria

*Primary case definition:*

- Presence of *P. falciparum* parasites density >5000 asexual forms/µL;

- AND one of more of the following criteria of disease severity:
    - Prostration
    - Respiratory distress
    - Blantyre coma score ≤ 2
    - Seizures: 2 or more
    - Hypoglycaemia < 2·2 mmol/L
    - Acidosis BE ≤-10·0 mmol/L
    - Lactate ≥ 5·0 mmol/L
    - Anaemia < 5·0 g/dL
- Without any of the following criteria of co-morbidity:
    - Pneumonia (confirmed by X-ray)
    - Meningitis (confirmed by cerebrospinal fluid examination)
    - Sepsis (with positive blood culture)
    - Gastroenteritis with dehydration

*Secondary case definitions:*

- Presence of *P. falciparum* asexual parasitaemia >5000 parasites/µL
    - AND one of the criteria of disease severity mentioned above
    - AND with diagnosis of a co-morbidity
- Presence of *P. falciparum* asexual parasitaemia >0 parasites/µL

  - AND one of the criteria of disease severity mentioned above
  - AND without diagnosis of a co-morbidity

##### 2.8.5 Efficacy against anaemia

*Incident severe anaemia*

Primary case definition:

- Documented haemoglobin (Hb) <5·0 g/dL identified at clinical presentation

Secondary case definitions:

- Documented Hb <5·0 g/dL identified at clinical presentation in association with *P. falciparum* asexual parasitaemia >0 parasites/µL
- Documented Hb <5·0 g/dL identified at clinical presentation and requirement for a blood transfusion
- Documented Hb <5·0 g/dL identified at clinical presentation in association with *P. falciparum* asexual parasitaemia > 5000 parasites/µL

*Prevalent anaemia*

Prevalent severe anaemia defined as:

- Documented Hb <5·0 g/dL identified

Prevalent moderate anaemia defined as:

- Documented Hb <8·0 g/dL

##### 2.8.6 Safety

The specific endpoints for safety and reactogenicity were actively and passively collected data on adverse events (AEs). The following parameters were assessed:

- Occurrence of solicited local reactogenicity signs and symptoms for 7 days following each vaccination (day of vaccination and 6 subsequent days)
- Occurrence of solicited systemic reactogenicity signs and symptoms for 7 days following each vaccination (day of vaccination and 6 subsequent days)
- Occurrence of unsolicited AEs for 28 days following the vaccination (day of vaccination and 27 subsequent days)
- Clinically significant change from baseline for safety laboratory measures throughout the study
- Occurrence of serious adverse events (SAEs) during the whole study duration

##### 2.8.7 Immunogenicity

Immunogenicity will be assessed by a variety of immunological assays, with comparison before and after vaccination. The following measures will be assessed:

- Serum ELISA response:
    - Quantitative antigen-specific IgG antibody levels (µg/mL readout) over time – analysis of peak responses and longevity;
    - Antigen-specific antibody subclass/isotype measurement;
    - Antigen-specific antibody avidity measurement;

- *In vitro* GIA against 3D7 clone *P. falciparum* parasites using purified total IgG and a single-cycle parasite lactate dehydrogenase (pLDH) readout assay
- Purified IgG ELISA versus GIA titration "Quality Analysis"

Available data are reported in this paper for Groups 1-4, including serum IgG ELISA (µg/mL readout) and *in vitro* GIA assay data, measured from all samples taken at baseline (screening) and 14 days post-third vaccination. All other immunological readouts will be reported at a later date.

##### 2.9 Vaccines

The design, production, preclinical and phase 1a/b testing of RH5.1 has been reported previously in detail.[1,2,7-9] Briefly, RH5.1 is a soluble protein immunogen based on the full-length *P. falciparum* reticulocyte-binding protein homologue 5 (RH5). The protein is produced by secretion from *Drosophila melanogaster* Schneider-2 (S2) cells provided by ExpreS$^2$ion Biotechnologies (Denmark). RH5.1 was developed at the University of Oxford and an initial batch, produced to Good Manufacturing Practice (GMP) by the Clinical Biomanufacturing Facility in Oxford,[1] was used in previously reported clinical trials VAC063 and VAC080.[2,9] Using the same established manufacturing processes, a second clinical batch of RH5.1 was filled in 2021 under GMP by a Contract Manufacturing Organisation based in the UK. This new batch of vaccine was used in the VAC091 phase 2b trial reported here.

Matrix-M adjuvant was manufactured in part by Novavax AB (Uppsala, Sweden) who produced the Matrix-A and -C fractions. These were then provided to the Serum Institute of India Pvt. Ltd. (Pune, India) [SIIPL] who manufactured the final Matrix-M adjuvant and provided it for the VAC091 trial. It is a potent, saponin-based adjuvant and comprises of partially-purified extracts of the bark of the *Quillaja saponaria* Molina tree, phosphatidylcholine and cholesterol. Matrix-M has demonstrated a favourable safety profile, as well as enhancement of cellular and humoral immune responses to a range of vaccine candidates and is licensed for human use in other malaria and Covid-19 vaccines.[4,10]

Rabivax-S was manufactured and supplied by SIIPL. It is an inactivated, freeze-dried, single-dose rabies vaccine. Rabivax-S was prequalified by the World Health Organisation in 2018.[11]

#### 2.10 Peripheral Blood Mononuclear Cell (PBMC), Plasma and Serum Preparation

Blood samples were collected into EDTA-treated vacutainer blood collection systems (Becton Dickinson, UK). PBMC were isolated and cryopreserved in foetal calf serum (FCS) containing 10% dimethyl sulfoxide and stored at -150°C within 6 h as previously described.[12] Plasma samples were stored at -70°C. For serum preparation, using untreated vacutainers, blood samples were stored at room temperature and then the clotted blood was centrifuged for 5 min (1000 *xg*). Serum was stored at -70°C.

#### 2.11 Immunological Analyses

##### 2.11.1 RH5 ELISA

Anti-RH5 total IgG ELISAs were performed against full-length RH5 protein (RH5.1) using standardised methodology as previously described.[9,13,14] The reciprocal of the test sample dilution giving an $OD_{405}$ of 1·0 in the standardised assay was used to assign an ELISA unit value of the standard. A standard curve and Gen5 ELISA software v3·04 (BioTek, UK) were used to convert the $OD_{405}$ of individual test samples into AU. These responses in AU are reported in µg/mL following generation of a conversion factor by calibration-free concentration analysis as reported previously.[13]

##### 2.11.2 Assay of Growth Inhibition Activity (GIA)

Standardised assays were performed by the GIA Reference Center, NIH, USA, using previously described methodology.[15] Researchers performing GIA assays were blinded to participant group allocation and timepoint. First, the full sample set (n=333) was tested in the order of volunteer ID using multiple batches of red blood cells (RBC). To minimise RBC batch effect[16] and the other assay-to-assay variation, samples that showed >19 % inhibition (considered as "GIA-positive" samples, n=222) in the initial GIA assay were randomly divided into 3 subgroups regardless of the volunteer ID, and the 3 subgroups were tested in three assays but using the same batch of RBC. For both tests, each sample was tested in triplicate wells in a single assay. For each assay, in brief, protein G purified IgG samples were incubated with RBC infected with synchronised *P. falciparum* 3D7 clone parasites in a final volume of 40 µL for 40 h at 37°C, and the final parasitaemia in each well was quantified by biochemical determination of pLDH.[9,14,16] All purified IgG samples were tested at 2·5 mg/mL final test well concentration.

### 3 Supplementary Figures and Tables

#### 3.1 **Table S1: VAC091 Study Groups.**

The primary vaccination schedules for Groups 1-4 only are shown. N indicates intended group size per protocol. Rabies = Rabivax-S. All vaccinations were given by the intramuscular route into the thigh.

| Group | N | Month 0 (Day 0) | Month 1 (Day 28) | Month 2 (Day 56) | Month 5 (Day 152) | Description |
|---|---|---|---|---|---|---|
| **1** | 60 | Rabies | Rabies | - | Rabies | Delayed third dose control |
| **2** | 120 | 10 µg RH5.1 / 50 µg Matrix-M | 10 µg RH5.1 / 50 µg Matrix-M | - | 10 µg RH5.1 / 50 µg Matrix-M | Delayed third dose regimen |
| **3** | 60 | Rabies | Rabies | Rabies | - | Monthly third dose control |
| **4** | 120 | 10 µg RH5.1 / 50 µg Matrix-M | 10 µg RH5.1 / 50 µg Matrix-M | 10 µg RH5.1 / 50 µg Matrix-M | - | Monthly third dose regimen |

#### 3.2 Table S2: Solicited AE Grading Scale.

| Adverse Event | Grade | Description |
|---|---|---|
| Pain at injection site | 0<br>1<br>2<br>3 | Absent<br>Minor reaction to touch<br>Cries/protests on touch<br>Cries when limb is moved/spontaneously painful |
| Swelling at injection site<br>*Record greatest surface diameter* | 0<br>1<br>2<br>3 | ≤3 mm<br>>3 - ≤20 mm<br>>20 - ≤50 mm<br>>50 mm |
| Redness/discoloration at injection site<br>*Record greatest surface diameter* | 0<br>1<br>2<br>3 | ≤3 mm<br>>3 - ≤20 mm<br>>20 - ≤50 mm<br>>50 mm |
| Fever | 0<br>1<br>2<br>3 | <37·5 °C<br>37·5-38·0 °C<br>>38·0-39·0 °C<br>>39·0 °C |
| Irritability/Fussiness | 0<br>1<br>2<br>3 | Behaviour as usual<br>Crying more than usual, no effect on normal activity<br>Crying more than usual, interferes with normal activity<br>Crying that cannot be comforted, prevents normal activity |
| Drowsiness | 0<br>1<br>2<br>3 | Behaviour as usual<br>Drowsiness easily tolerated<br>Drowsiness that interferes with normal activity<br>Drowsiness that prevents normal activity |
| Loss of appetite | 0<br>1<br>2<br>3 | Appetite as usual<br>Eating less than usual, no effect on normal activity<br>Eating less than usual, interferes with normal activity<br>Not eating at all |

##### 3.3 Table S3: General severity grading criteria for AEs.

| Grade | Severity |
|---|---|
| GRADE 0 | None |
| GRADE 1 | Mild: Transient or mild discomfort (<48 hours); no medical intervention/therapy required |
| GRADE 2 | Moderate: Mild to moderate limitation in activity – some assistance may be needed; no or minimal medical intervention/therapy required |
| GRADE 3 | Severe: Marked limitation in activity, some assistance usually required; may require medical intervention/therapy |

#### 3.4 Table S4: Schedule of Events for Delayed Third Dose Regimen Groups (VAC091 Groups 1-2).

Schedule shown up to 6 months post-third vaccination.

| | Screening | Vacc 1 | | | Vacc 2 | | | | | | Vacc 3 | | | | | | | | |
|---|---|---|---|---|---|---|---|---|---|---|---|---|---|---|---|---|---|---|---|
| **Study visit** | **1** | **2** | **3-7** | **8** | **9** | **10-15** | **16** | **17** | **18** | **19** | **20** | **21-26** | **27** | **28** | **29** | **30** | **31** | **32** | **33** |
| **Day of visit** | -30 to D0 | D0 | D1-6 | D14 | D28 | D29-34 | D42 | D56 | D86 | D116 | D152 | D153-158 | D166 | D180 | D210 | D240 | D270 | D300 | D332 |
| **Window period** | -30 | 0 | 0 | ±2 | -7/+14 | 0 | ±2 | -7/14 | ±14 | ±14 | -7/+14 | 0 | ±2 | -7/14 | ±14 | ±14 | ±14 | ±14 | ±14 |
| **Clinic visit** | X | X | | X | X | | X | X | | | X | | X | X | X | | | | X |
| **Home visit** | | | X | | | X | | | X | X | | X | | | | X | X | X | |
| **STUDY procedures** | | | | | | | | | | | | | | | | | | | |
| **Issue of identification card** | X | | | | | | | | | | | | | | | | | | |
| **Check of identification card** | | X | X | X | X | X | X | X | X | X | X | X | X | X | X | X | X | X | X |
| **Informed consent** | X | | | | | | | | | | | | | | | | | | |
| **Inclusion/Exclusion criteria** | X | X | | | X | | | | | | X | | | | | | | | |
| **Demographics** | X | | | | | | | | | | | | | | | | | | |
| **Medical history** | X | X | | X | X | | X | X | | | X | | X | X | X | | | | X |
| **Physical Examination** | X | (X) | | (X) | (X) | | (X) | (X) | | | (X) | | (X) | (X) | (X) | | | | (X) |
| **Anthropometry** | X | X | | | | | X | | | | X | | X | | X | | | | X |
| **Temperature** | | X | X | X | X | X | X | X | X | X | X | X | X | X | X | X | X | X | X |
| **Randomisation** | | X | | | | | | | | | | | | | | | | | |
| **Vaccination** | | X | | | X | | | | | | X | | | | | | | | |
| **Recording of concomitant medication** | X | X | X | X | X | X | X | X | X | X | X | X | X | X | X | X | X | X | X |
| **Recording of use of SMC** | X | X | | | X | | | X | X | X | X | | | X | X | X | X | X | X |
| **Recording of EPI/ other vaccine doses** | X | X | | | X | | | X | | | X | | | X | | | | | X |
| **Feeding history** | X | | | | X | | | X | X | X | X | | | X | X | X | X | X | X |
| **Documentation of ITN and IRS** | X | | | | X | | | X | X | X | X | | | X | X | X | X | X | X |
| **Recording of covariates** | X | | | | | | | | | | | | | | | | | | |
| **Photographs** | X | (X) | | (X) | (X) | | (X) | (X) | | | (X) | | (X) | (X) | (X) | | | | (X) |

| | Screening | Vacc 1 | | | Vacc 2 | | | | | | Vacc 3 | | | | | | | | |
|---|---|---|---|---|---|---|---|---|---|---|---|---|---|---|---|---|---|---|---|
| **Study visit** | **1** | **2** | **3-7** | **8** | **9** | **10-15** | **16** | **17** | **18** | **19** | **20** | **21-26** | **27** | **28** | **29** | **30** | **31** | **32** | **33** |
| **Day of visit** | -30 to D0 | D0 | D1-6 | D14 | D28 | D29-34 | D42 | D56 | D86 | D116 | D152 | D153-158 | D166 | D180 | D210 | D240 | D270 | D300 | D332 |
| **Clinic visit** | X | X | | X | X | | X | X | | | X | | X | X | X | | | | X |
| **Home visit** | | | X | | | X | | | X | X | | X | | | | X | X | X | |
| **SAFETY procedures** | | | | | | | | | | | | | | | | | | | |
| **Recording of solicited AEs** | | X | X | | X | X | | | | | X | X | | | | | | | |
| **Recording of unsolicited AEs** | | X | X | X | X | X | X | | | | X | X | X | X | | | | | |
| **Recording of SAEs** | | X | X | X | X | X | X | X | X | X | X | X | X | X | X | X | X | X | X |
| **BLOOD collection** | | | | | | | | | | | | | | | | | | | |
| **Blood film for Plasmodium species** | | X | | | | | | | | | | | | | | | | | X |
| **Dried blood spot** | | X | | X | X | | X | X | X | X | X | | X | X | X | X | X | X | X |
| **Blood film for Plasmodium species if temp** ≥37.5°C or fever within 24 hours | | | | X | X | | X | X | X | X | X | | X | X | X | X | X | X | |
| **Haematology & Biochemistry (mL)** | 1.5 | | | | | | 1.5 | | | | 1.5 | | | | 1.5 | | | | 1.5 |
| **Immunology (mL)** | 5 | 5 | | | | | 5 | 5 | | | 5 | | 5 | | 5 | | | | 5 |
| **Total blood volume (mL)** | 6.5 | 5 | 0 | 0 | 0 | 0 | 6.5 | 5 | 0 | 0 | 6.5 | 0 | 5 | 0 | 6.5 | 0 | 0 | 0 | 6.5 |
| **Cumulative blood volume (mL)** | 6.5 | 11.5 | 11.5 | 11.5 | 11.5 | 11.5 | 18 | 23 | 23 | 23 | 29.5 | 29.5 | 34.5 | 34.5 | 41 | 41 | 41 | 41 | 47.5 |

##### 3.5 Table S5: Schedule of Events for Monthly Third Dose Regimen Groups (VAC091 Groups 3-4).

Schedule shown up to 6 months post-third vaccination.

| | Screening | Vacc 1 | | | Vacc 2 | | | Vacc 3 | | | | | | | | |
|---|---|---|---|---|---|---|---|---|---|---|---|---|---|---|---|---|
| **Study visit** | **1** | **2** | **2-7** | **8** | **9** | **10-15** | **16** | **17** | **18-23** | **24** | **25** | **26** | **27** | **28** | **29** | **30** |
| **Day of visit** | -30 to D0 | D0 | D1-6 | D14 | D28 | D29-34 | D42 | D56 | D57-62 | D70 | D84 | D114 | D144 | D174 | D204 | D238 |
| **Window period** | -30 | 0 | 0 | ±2 | -7/+14 | 0 | ±2 | -7/+14 | 0 | ±2 | -7/14 | ±14 | ±14 | ±14 | ±14 | ±14 |
| **Clinic visit** | X | X | | X | X | | X | X | | X | X | X | | | | X |
| **Home visit** | | | X | | | X | | | X | | | | X | X | X | |
| **STUDY procedures** | | | | | | | | | | | | | | | | |
| **Issue of identification card** | X | | | | | | | | | | | | | | | |
| **Check of identification card** | | X | X | X | X | X | X | X | X | X | X | X | X | X | X | X |
| **Informed consent** | X | | | | | | | | | | | | | | | |
| **Inclusion/Exclusion criteria** | X | X | | | X | | | X | | | | | | | | |
| **Demographics** | X | | | | | | | | | | | | | | | |
| **Medical history** | X | X | | X | X | | X | X | | X | X | X | | | | X |
| **Physical Examination** | X | (X) | | (X) | (X) | | (X) | (X) | | (X) | (X) | (X) | | | | (X) |
| **Anthropometry** | X | X | | | | | X | X | | X | | X | | | | X |
| **Temperature** | | X | X | X | X | X | X | X | X | X | X | X | X | X | X | X |
| **Randomisation** | | X | | | | | | | | | | | | | | |
| **Vaccination** | | X | | | X | | | X | | | | | | | | |
| **Recording of concomitant medication** | X | X | X | X | X | X | X | X | X | X | X | X | X | X | X | X |
| **Recording of use of SMC** | X | X | | | X | | | X | | | X | X | X | X | X | X |
| **Recording of EPI/ other vaccine doses** | X | X | | | X | | | X | | | X | | | | | X |
| **Feeding history** | X | | | | X | | | X | | | X | X | X | X | X | X |
| **Documentation of ITN and IRS** | X | | | | X | | | X | | | X | X | X | X | X | X |
| **Recording of covariates** | X | | | | | | | | | | | | | | | |
| **Photographs** | X | (X) | | (X) | (X) | | (X) | (X) | | (X) | (X) | (X) | | | | (X) |

| | Screening | Vacc 1 | | | Vacc 2 | | | Vacc 3 | | | | | | | | |
|---|---|---|---|---|---|---|---|---|---|---|---|---|---|---|---|---|
| **Study visit** | **1** | **2** | **2-7** | **8** | **9** | **10-15** | **16** | **17** | **18-23** | **24** | **25** | **26** | **27** | **28** | **29** | **30** |
| **Day of visit** | -30 to D0 | D0 | D1-6 | D14 | D28 | D29-34 | D42 | D56 | D57-62 | D70 | D84 | D114 | D144 | D174 | D204 | D238 |
| **SAFETY procedures** | | | | | | | | | | | | | | | | |
| **Recording of solicited AEs** | | X | X | | X | X | | X | X | | | | | | | |
| **Recording of unsolicited AEs** | | X | X | X | X | X | X | X | X | X | X | | | | | |
| **Recording of SAEs** | | X | X | X | X | X | X | X | X | X | X | X | X | X | X | X |
| **BLOOD collection** | | | | | | | | | | | | | | | | |
| **Blood film for Plasmodium species** | | X | | | | | | | | | | | | | | X |
| **Dried blood spots** | | X | | X | X | | X | X | | X | X | X | X | X | X | X |
| **Blood film for Plasmodium species if temp** ≥37.5°C or fever within 24 hours | | | | X | X | | X | X | | X | X | X | X | X | X | |
| **Haematology & Biochemistry (mL)** | 1.5 | | | | | | 1.5 | | | | | 1.5 | | | | 1.5 |
| **Immunology (mL)** | 5 | 5 | | | | | 5 | 5 | | 5 | | 5 | | | | 5 |
| **Total blood volume (mL)** | 6.5 | 5 | 0 | 0 | 0 | 0 | 6.5 | 5 | 0 | 5 | 0 | 6.5 | 0 | 0 | 0 | 6.5 |
| **Cumulative blood volume (mL)** | 6.5 | 11.5 | 11.5 | 11.5 | 11.5 | 11.5 | 18 | 23 | 23 | 28 | 28 | 34.5 | 34.5 | 34.5 | 34.5 | 41 |

#### 3.6 Table S6: Comparability of trial sample at screening.

Table includes 338 trial children who received all eligible doses (3) of RH5.1/Matrix-M (MM) or Rabivax-S (rabies) vaccine. *SMC=seasonal malaria chemoprevention in 2023 (up to four monthly rounds of preventative malaria treatment); IRS = Indoor residual spraying with insecticide; ITN = insecticide treated net.

| **Characteristic** | **Overall** | **Delayed Rabies** | **Delayed RH5.1/MM** | **Monthly Rabies** | **Monthly RH5.1/MM** |
|---|---|---|---|---|---|
| **Age in months** (mean SD) | 10·5 (3·6) | 10·5 (3·7) | 10·6 (3·6) | 10·3 (3·6) | 10·5 (3·5) |
| **Age category in months** (n %) | | | | | |
| 5-8 months | 117 (34·6) | 19 (34·6) | 36 (31·6) | 21 (36·8) | 41 (36·6) |
| 9-12 months | 92 (27·2) | 16 (29·1) | 35 (30·7) | 13 (22·8) | 28 (25·0) |
| 13-17 months | 129 (38·2) | 20 (36·4) | 43 (37·7) | 23 (40·4) | 43 (38·4) |
| **Female** (n %) | 177 (52·4) | 29 (52·7) | 59 (51·8) | 29 (50·9) | 60 (53·6) |
| **ITN* use the night before** (n %) | | | | | |
| Yes - adequate | 290 (85·8) | 43 (78·2) | 86 (75·4) | 52 (91·2) | 109 (97·3) |
| Yes – ITN damaged | 7 (2·1) | 1 (1·8) | 6 (5·3) | 0 (0·0) | 0 (0·0) |
| No – has ITN | 27 (8·0) | 9 (16·4) | 14 (12·3) | 3 (5·3) | 1 (0·9) |
| No – no ITN | 14 (4·1) | 2 (3·6) | 8 (7·0) | 2 (3·5) | 2 (1·8) |
| **IRS* in the past year** (n %) | 8 (2·4) | 4 (7·3) | 4 (3·5) | 0 (0·0) | 0 (0·0) |
| **SMC* coverage** (n %) | | | | | |
| At least 1 round | 333 (98·5) | 54 (98·2) | 111 (97·4) | 57 (100·0) | 111 (99·1) |
| 1 round | 33 (7·8) | 5 (9·1) | 23 (20·2) | 2 (3·5) | 3 (2·7) |
| 2 rounds | 72 (21·3) | 13 (23·6) | 28 (24·6) | 12 (21·1) | 19 (17·0) |
| 3 rounds | 116 (34·3) | 23 (41·8) | 27 (23·7) | 20 (35·1) | 46 (41·1) |
| 4 rounds | 112 (33·1) | 13 (23·6) | 33 (29·0) | 23 (40·4) | 43 (38·4) |
| Missing SMC info | 5 (1·5) | 1 (1·8) | 3 (2·6) | 0 (0·0) | 1 (0·9) |
| **Weight for age z scores** (n %) | | | | | |
| -3 to -2 | 38 (11·2) | 8 (14·6) | 18 (15·8) | 6 (10·5) | 6 (5·4) |
| -2 to -1 | 94 (27·8) | 13 (23·6) | 29 (25·4) | 14 (24·6) | 38 (33·9) |
| -1SD to Median | 141 (41·7) | 21 (38·2) | 46 (40·4) | 27 (47·4) | 47 (42·0) |
| Median to +1 | 51 (15·1) | 12 (21·8) | 15 (13·2) | 8 (14·0) | 16 (14·3) |
| +1 to +2 | 13 (3·9) | 1 (1·8) | 6 (5·3) | 1 (1·8) | 5 (4·5) |
| +2 to +3 | 1 (0·3) | 0 (0·0) | 0 (0·0) | 1 (1·8) | 0 (0·0) |

##### 3.7 Table S7: Per-protocol sample additional endpoint at 6 months: Analysis of time to first episode of clinical malaria comparing between the two control groups.

Cox regression models were used to analyse time to first episode of clinical malaria (presence of axillary temperature ≥37·5°C and/or history of fever within the last 24 hours, *P. falciparum* asexual parasitaemia concentration according to case definition) between the two control groups. Vaccine Efficacy = 1 – hazard ratio. 95% CI = 95% Confidence Interval. 6 month analysis timepoint and per-protocol sample. Control group data are combined in the trial impact analysis as no differences were observed in the clinical malaria endpoint between the two control groups at any parasite concentration. * Adjusted vaccine efficacy: includes covariates of total number of rounds of SMC received, ITN use (adequate or not) the night before screening visit, and age categories 5-8 months, 9-12 months, 13-17 months. Excludes 1 child with missing data on covariates in the Delayed Rabies arm. ** Event rate per 100 days (where event is the first episode of clinical malaria) is also reported for information only, calculated as: (the number of events / [child days/100]).

| **Groups** | **N** | **Event rate per 100 days **** | **VACCINE EFFICACY (95% CI)** | **p** | **VACCINE EFFICACY (95% CI) [adjusted*]** | **p [adjusted]** |
|---|---|---|---|---|---|---|
| **Primary Case Definition** (parasitaemia >**5000** parasites/μL) | | | | | | |
| Delayed Rabies | 55 | 0·23 (17/74·8) | 1 | | 1 | |
| Monthly Rabies | 57 | 0·20 (16/81·4) | 0·14 (-0·70, 0·56) | 0·6675 | 0·20 (-0·62, 0·60) | 0·5399 |
| **Secondary Case Definition** (parasitaemia >**0** parasites/μL) | | | | | | |
| Delayed Rabies | 55 | 0·31 (21/68·5) | 1 | | 1 | |
| Monthly Rabies | 57 | 0·23 (18/78·9) | 0·24 (-0·43, 0·59) | 0·3995 | 0·28 (-0·38, 0·63) | 0·3216 |
| **Secondary Case Definition** (parasitaemia >**20,000** parasites/μL) | | | | | | |
| Delayed Rabies | 55 | 0·18 (14/80·0) | 1 | | 1 | |
| Monthly Rabies | 57 | 0·17 (14/84·4) | 0·05 (-0·99, 0·55) | 0·8846 | 0·07 (-0·98, 0·57) | 0·8462 |
| **Additional Case Definition** (parasitaemia >**50,000** parasites/μL) | | | | | | |
| Delayed Rabies | 55 | 0·12 (10/86·3) | 1 | | 1 | |
| Monthly Rabies | 57 | 0·10 (9/92·4) | 0·16 (-1·06, 0·66) | 0·6951 | 0·26 (-0·85, 0·71) | 0·5191 |
| **Additional Case Definition** (parasitaemia >**100,000** parasites/μL) | | | | | | |
| Delayed Rabies | 55 | 0·05 (5/93·7) | 1 | | 1 | |
| Monthly Rabies | 57 | 0·06 (6/96·5) | -0·16 (-2·79, 0·65) | 0·8085 | -0·02 (-2·37, 0·69) | 0·9787 |

#### 3.8 Table S8: Per-protocol sample additional endpoint at 6 months: Analysis of time to first episode of clinical malaria comparing between the two RH5.1/MM regimens.

Cox regression models were used to analyse time to first episode of clinical malaria (presence of axillary temperature ≥37·5°C and/or history of fever within the last 24 hours, *P. falciparum* asexual parasitaemia concentration according to case definition) between the two RH5.1/MM regimens. Vaccine Efficacy = 1 – hazard ratio. 95% CI = 95% Confidence Interval. 6 month analysis timepoint and per-protocol sample.

* Adjusted vaccine efficacy: includes covariates of total number of rounds of SMC received, ITN use (adequate or not) the night before screening visit, and age categories 5-8 months, 9-12 months, 13-17 months. Excludes 4 children with missing data on covariates: 3 in the Delayed RH5.1/MM arm and 1 in the Monthly RH5.1/MM arm. ** Event rate per 100 days (where event is the first episode of clinical malaria) is also reported for information only, calculated as: (the number of events / [child days/100]).

| Groups | N | Event rate per 100 days ** | VACCINE EFFICACY (95% CI) | p | VACCINE EFFICACY (95% CI) [adjusted*] | p [adjusted] |
|---|---|---|---|---|---|---|
| **Primary Case Definition** (parasitaemia >**5000** parasites/µL) | | | | | | |
| Monthly RH5.1/MM | 112 | 0·12 (22/176·4) | 1 | | 1 | |
| Delayed RH5.1/MM | 114 | 0·09 (17/185·0) | 0·26 (-0·39, 0·61) | 0·3464 | 0·33 (-0·35, 0·66) | 0·2671 |
| **Secondary Case Definition** (parasitaemia >**0** parasites/µL) | | | | | | |
| Monthly RH5.1/MM | 112 | 0·25 (39/158·9) | 1 | | 1 | |
| Delayed RH5.1/MM | 114 | 0·15 (26/176·5) | 0·40 (0·01, 0·63) | 0·0438 | 0·44 (0·02, 0·68) | 0·0419 |
| **Secondary Case Definition** (parasitaemia >**20,000** parasites/µL) | | | | | | |
| Monthly RH5.1/MM | 112 | 0·09 (16/183·0) | 1 | | 1 | |
| Delayed RH5.1/MM | 114 | 0·05 (10/194·5) | 0·40 (-0·31, 0·73) | 0·1991 | 0·25 (-0·71, 0·67) | 0·4905 |
| **Additional Case Definition** (parasitaemia >**50,000** parasites/µL) | | | | | | |
| Monthly RH5.1/MM | 112 | 0·01 (2/201·9) | 1 | | 1 | |
| Delayed RH5.1/MM | 114 | 0·02 (4/203·4) | -0·97 (-9·77, 0·64) | 0·4325 | -1·74 (-14·55, 0·52) | 0·2549 |
| **Additional Case Definition** (parasitaemia >**100,000** parasites/µL) | | | | | | |
| Monthly RH5.1/MM | 112 | 0·00 (1/203·1) | 1 | | 1 | |
| Delayed RH5.1/MM | 114 | 0·01 (2/206·3) | -0·96 (-20·6, 0·82) | 0·583 | -1·35 (-26·45, 0·8) | 0·4946 |

##### 3.9 Table S9. Per-protocol sample additional endpoint at 6 months: Analysis of vaccine efficacy against all clinical malaria episodes.

A secondary analysis of vaccine efficacy against all clinical malaria episodes was carried out, using Cox regression models with a robust standard error to account for multiple episodes. Episodes occurring within 14 days of a previous episode were classed as the same event. For participants without an episode of clinical malaria, their time was censored at the date of their withdrawal or the date of their 6-month blood sampling (noting no deaths have occurred in this trial). Clinical malaria was defined as presence of axillary temperature ≥37·5°C and/or history of fever within the last 24 hours, *P. falciparum* asexual parasitaemia concentration according to case definition. Vaccine Efficacy = 1 – hazard ratio. 95% CI = 95% Confidence Interval. 6 month analysis timepoint and per-protocol sample. * Adjusted vaccine efficacy: includes covariates of total number of rounds of SMC received, ITN use (adequate or not) the night before screening visit, and age categories 5-8 months, 9-12 months, 13-17 months. Excludes 5 children with missing data on covariates: 1 in the Delayed Rabies arm, 3 in the Delayed RH5.1/MM arm and 1 in the Monthly RH5.1/MM arm. ** Event rate per 100 days (where event is all episodes of clinical malaria) is also reported for information only, calculated as: (the number of events / [child days/100]).

| Groups | N | Number of malaria episodes per child n (%) | | | | Event rate per 100 days ** | VACCINE EFFICACY (95% CI) | p | VACCINE EFFICACY (95% CI) [adjusted*] | p [adjusted] |
|---|---|---|---|---|---|---|---|---|---|---|
| | | 0 | 1 | 2 | 3 | | | | | |
| **Primary Case Definition** (parasitaemia >**5000** parasites/µL) | | | | | | | | | | |
| Rabies controls (combined) | 112 | 79 (70·5) | 26 (23·2) | 7 (6·3) | 0 (0·0) | 0·19 (40/204·6) | 1 | | 1 | |
| Delayed RH5.1/MM | 114 | 97 (85·1) | 16 (14·0) | 1 (0·9) | 0 (0·0) | 0·08 (18/208·3) | 0·56 (0·24, 0·74) | 0·0035 | 0·56 (0·25, 0·75) | 0·0026 |
| Monthly RH5.1/MM | 112 | 90 (80·4) | 20 (17·9) | 2 (1·8) | 0 (0·0) | 0·11 (24/204·6) | 0·40 (0·01, 0·64) | 0·0446 | 0·41 (0·04, 0·64) | 0·0348 |
| **Secondary Case Definition** (parasitaemia >**0** parasites/µL) | | | | | | | | | | |
| Rabies controls (combined) | 112 | 73 (65·2) | 32 (28·6) | 6 (5·4) | 1 (0·9) | 0·22 (47/204·6) | 1 | | 1 | |
| Delayed RH5.1/MM | 114 | 88 (77·2) | 25 (21·9) | 1 (0·9) | 0 (0·0) | 0·13 (27/208·3) | 0·44 (0·12, 0·64) | 0·0119 | 0·42 (0·11, 0·63) | 0·0140 |
| Monthly RH5.1/MM | 112 | 73 (65·2) | 36 (32·1) | 3 (2·7) | 0 (0·0) | 0·20 (42/204·6) | 0·11 (-0·32, 0·39) | 0·5685 | 0·10 (-0·34, 0·39) | 0·6130 |
| **Secondary Case Definition** (parasitaemia >**20,000** parasites/µL) | | | | | | | | | | |
| Rabies controls (combined) | 112 | 84 (75·0) | 22 (19·6) | 6 (5·4) | 0 (0·0) | 0·16 (34/204·6) | 1 | | 1 | |
| Delayed RH5.1/MM | 114 | 104 (91·2) | 10 (8·8) | 0 (0·0) | 0 (0·0) | 0·05 (10/208·3) | 0·71 (0·43, 0·85) | 0·0004 | 0·69 (0·38, 0·85) | 0·0009 |
| Monthly RH5.1/MM | 112 | 96 (85·7) | 14 (12·5) | 2 (1·8) | 0 (0·0) | 0·08 (18/204·6) | 0·47 (0·05, 0·71) | 0·0341 | 0·50 (0·12, 0·71) | 0·0156 |
| **Additional Case Definition** (parasitaemia >**50,000** parasites/µL) | | | | | | | | | | |
| Rabies controls (combined) | 112 | 93 (83) | 15 (13·4) | 4 (3·6) | 0 (0·0) | 0·11 (23/204·6) | 1 | | 1 | |
| Delayed RH5.1/MM | 114 | 110 (96·5) | 4 (3·5) | 0 (0·0) | 0 (0·0) | 0·02 (4/208·3) | 0·83 (0·51, 0·94) | 0·0011 | 0·81 (0·43, 0·94) | 0·0032 |
| Monthly RH5.1/MM | 112 | 110 (98·2) | 2 (1·8) | 0 (0·0) | 0 (0·0) | 0·01 (2/204·6) | 0·91 (0·63, 0·98) | 0·0009 | 0·92 (0·66, 0·98) | 0·0007 |
| **Additional Case Definition** (parasitaemia >**100,000** parasites/µL) | | | | | | | | | | |
| Rabies controls (combined) | 112 | 101 (90·2) | 11 (9·8) | 0 (0·0) | 0 (0·0) | 0·05 (11/204·6) | 1 | | 1 | |
| Delayed RH5.1/MM | 114 | 112 (98·3) | 2 (1·8) | 0 (0·0) | 0 (0·0) | 0·01 (2/208·3) | 0·82 (0·21, 0·96) | 0·0231 | 0·81 (0·09, 0·96) | 0·0380 |
| Monthly RH5.1/MM | 112 | 111 (99·1) | 1 (0·9) | 0 (0·0) | 0 (0·0) | 0·00 (1/204·6) | 0·91 (0·31, 0·99) | 0·0208 | 0·91 (0·33, 0·99) | 0·0190 |

##### 3.10 Table S10. Per-protocol sample additional endpoint at 6 months: Analysis of vaccine efficacy against all clinical malaria episodes comparing between the two RH5.1/Matrix-M regimens.

A secondary analysis of vaccine efficacy against all clinical malaria episodes comparing between the RH5.1/Matrix-M regimens was carried out, using Cox regression models with a robust standard error to account for multiple episodes. Episodes occurring within 14 days of a previous episode were classed as the same event. For participants without an episode of clinical malaria, their time was censored at the date of their withdrawal or the date of their 6-month blood sampling (noting no deaths have occurred in this trial). Clinical malaria was defined as presence of axillary temperature ≥37·5°C and/or history of fever within the last 24 hours, *P. falciparum* asexual parasitaemia concentration according to case definition. Vaccine Efficacy = 1 – hazard ratio. 95% CI = 95% Confidence Interval. 6 month analysis timepoint and per-protocol sample. * Adjusted vaccine efficacy: includes covariates of total number of rounds of SMC received, ITN use (adequate or not) the night before screening visit, and age categories 5-8 months, 9-12 months, 13-17 months. Excludes 4 children with missing data on covariates: 3 in the Delayed RH5.1/MM arm and 1 in the Monthly RH5.1/MM arm. ** Event rate per 100 days (where event is all episodes of clinical malaria) is also reported for information only, calculated as: (the number of events / [child days/100]).

| **Groups** | **N** | **Number of malaria episodes per child n (%)** | | | | **Event rate per 100 days **** | **VACCINE EFFICACY (95% CI)** | **p** | **VACCINE EFFICACY (95% CI) [adjusted*]** | **p [adjusted]** |
|---|---|---|---|---|---|---|---|---|---|---|
| | | **0** | **1** | **2** | **3** | | | | | |
| **Primary Case Definition** (parasitaemia >**5000** parasites/µL) | | | | | | | | | | |
| Monthly RH5.1/MM | 112 | 90 (80·4) | 20 (17·9) | 2 (1·8) | 0 (0·0) | 0·11 (24/204·6) | 1 | | 1 | |
| Delayed RH5.1/MM | 114 | 97 (85·1) | 16 (14·0) | 1 (0·9) | 0 (0·0) | 0·08 (18/208·3) | 0·26 (-0·34, 0·59) | 0·3168 | 0·33 (-0·32, 0·66) | 0·2496 |
| **Secondary Case Definition** (parasitaemia >**0** parasites/µL) | | | | | | | | | | |
| Monthly RH5.1/MM | 112 | 73 (65·2) | 36 (32·1) | 3 (2·7) | 0 (0·0) | 0·20 (42/204·6) | 1 | | 1 | |
| Delayed RH5.1/MM | 114 | 88 (77·2) | 25 (21·9) | 1 (0·9) | 0 (0·0) | 0·13 (27/208·3) | 0·37 (0·02, 0·59) | 0·0389 | 0·42 (0·04, 0·65) | 0·0326 |
| **Secondary Case Definition** (parasitaemia >**20,000** parasites/µL) | | | | | | | | | | |
| Monthly RH5.1/MM | 112 | 96 (85·7) | 14 (12·5) | 2 (1·8) | 0 (0·0) | 0·08 (18/204·6) | 1 | | 1 | |
| Delayed RH5.1/MM | 114 | 104 (91·2) | 10 (8·8) | 0 (0·0) | 0 (0·0) | 0·05 (10/208·3) | 0·45 (-0·17, 0·75) | 0·1190 | 0·34 (-0·52, 0·71) | 0·3329 |
| **Additional Case Definition** (parasitaemia >**50,000** parasites/µL) | | | | | | | | | | |
| Monthly RH5.1/MM | 112 | 110 (98·2) | 2 (1·8) | 0 (0·0) | 0 (0·0) | 0·01 (2/204·6) | 1 | | 1 | |
| Delayed RH5.1/MM | 114 | 110 (96·5) | 4 (3·5) | 0 (0·0) | 0 (0·0) | 0·02 (4/208·3) | -0·96 (-9·55, 0·63) | 0·4310 | -1·70 (-19·21, 0·64) | 0·3329 |
| **Additional Case Definition** (parasitaemia >**100,000** parasites/µL) | | | | | | | | | | |
| Monthly RH5.1/MM | 112 | 111 (99·1) | 1 (0·9) | 0 (0·0) | 0 (0·0) | 0·00 (1/204·6) | 1 | | 1 | |
| Delayed RH5.1/MM | 114 | 112 (98·3) | 2 (1·8) | 0 (0·0) | 0 (0·0) | 0·01 (2/208·3) | -0·96 (-20·48, 0·82) | 0·5799 | -1·36 (-42·41, 0·87) | 0·5643 |

#### 3.11 Table S11: Intention-to-treat (ITT) sample endpoint at 6 months: Analysis of time to first episode of clinical malaria.

Cox regression models were used to analyse the secondary endpoint of time to first episode of clinical malaria based on the ITT population at the 6-month timepoint. Clinical malaria is defined as measured axillary temperature ≥37·5°C and/or reported history of fever within the last 24 hours, and *P. falciparum* asexual parasitaemia concentration according to each case definition. Vaccine Efficacy = 1 – hazard ratio. 95% CI = 95% Confidence Interval. * Adjusted vaccine efficacy: includes covariates of total number of rounds of SMC received, ITN use (adequate or not) the night before the screening visit, and age categories 5-8 months, 9-12 months, 13-17 months. Excludes 5 children with missing data on covariates: 1 in the Delayed Rabies arm, 3 in the Delayed RH5.1/Matrix-M arm and 1 in the Monthly RH5.1/Matrix-M arm. ** Event rate per 100 days (where event is the first episode of clinical malaria) is also reported for information only, calculated as: (the number of events / [child days/100]).

| Groups | N | Event rate per 100 days ** | VACCINE EFFICACY (95% CI) | p | VACCINE EFFICACY (95% CI) [adjusted*] | p [adjusted] |
|---|---|---|---|---|---|---|
| **Primary Case Definition** (parasitaemia >**5000** parasites/µL) | | | | | | |
| Rabies controls (combined) | 119 | 0·21 (34/159·3) | 1 | | 1 | |
| Delayed RH5.1/MM | 118 | 0·09 (17/186·5) | 0·56 (0·22, 0·76) | 0·0054 | 0·56 (0·20, 0·76) | 0·0068 |
| Monthly RH5.1/MM | 117 | 0·12 (22/185·6) | 0·43 (0·02, 0·67) | 0·0408 | 0·43 (0·02, 0·67) | 0·0435 |
| **Secondary Case Definition** (parasitaemia >**0** parasites/µL) | | | | | | |
| Rabies controls (combined) | 119 | 0·27 (40/150·3) | 1 | | 1 | |
| Delayed RH5.1/MM | 118 | 0·15 (26/177·9) | 0·45 (0·10, 0·66) | 0·0183 | 0·42 (0·03, 0·65) | 0·0371 |
| Monthly RH5.1/MM | 117 | 0·23 (39/168·0) | 0·12 (-0·36, 0·44) | 0·5573 | 0·10 (-0·42, 0·43) | 0·6509 |
| **Secondary Case Definition** (parasitaemia >**20,000** parasites/µL) | | | | | | |
| Rabies controls (combined) | 119 | 0·17 (28/169·0) | 1 | | 1 | |
| Delayed RH5.1/MM | 118 | 0·05 (10/196·0) | 0·68 (0·34, 0·84) | 0·0020 | 0·67 (0·31, 0·84) | 0·0031 |
| Monthly RH5.1/MM | 117 | 0·08 (16/192·2) | 0·48 (0·04, 0·72) | 0·0365 | 0·53 (0·13, 0·75) | 0·0163 |
| **Additional Case Definition** (parasitaemia >**50,000** parasites/µL) | | | | | | |
| Rabies controls (combined) | 119 | 0·10 (19/183·3) | 1 | | 1 | |
| Delayed RH5.1/MM | 118 | 0·02 (4/204·9) | 0·81 (0·43, 0·93) | 0·0029 | 0·79 (0·37, 0·93) | 0·0053 |
| Monthly RH5.1/MM | 117 | 0·01 (2/211·0) | 0·90 (0·59, 0·98) | 0·0016 | 0·91 (0·63, 0·98) | 0·0010 |
| **Additional Case Definition** (parasitaemia >**100,000** parasites/µL) | | | | | | |
| Rabies controls (combined) | 119 | 0·06 (11/194·8) | 1 | | 1 | |
| Delayed RH5.1/MM | 118 | 0·01 (2/207·8) | 0·83 (0·22, 0·96) | 0·0223 | 0·82 (0·16, 0·96) | 0·0291 |
| Monthly RH5.1/MM | 117 | 0·00 (1/212·2) | 0·91 (0·34, 0·99) | 0·0180 | 0·92 (0·40, 0·99) | 0·0146 |

##### 3.12 Table S12: Intention-to-treat (ITT) sample endpoint at 6 months: Analysis of time to first episode of clinical malaria comparing between the two RH5.1/MM regimens.

Cox regression models were used to analyse time to first episode of clinical malaria (presence of axillary temperature ≥37·5°C and/or history of fever within the last 24 hours, *P. falciparum* asexual parasitaemia concentration according to case definition) comparing between the two RH5.1/MM regimens and based on the ITT population at the 6-month timepoint. Vaccine Efficacy = 1 – hazard ratio. 95% CI = 95% Confidence Interval. * Adjusted vaccine efficacy: includes covariates of total number of rounds of SMC received, ITN use (adequate or not) the night before screening visit, and age categories 5-8 months, 9-12 months, 13-17 months. Excludes 4 children with missing data on covariates: 3 in the Delayed RH5.1/MM arm and 1 in the Monthly RH5.1/MM arm. ** Event rate per 100 days (where event is the first episode of clinical malaria) is also reported for information only, calculated as: (the number of events / [child days/100]).

| Groups | N | Event rate per 100 days ** | VACCINE EFFICACY (95% CI) | p | VACCINE EFFICACY (95% CI) [adjusted*] | p [adjusted] |
|---|---|---|---|---|---|---|
| **Primary Case Definition** (parasitaemia >**5000** parasites/µL) | | | | | | |
| Monthly RH5.1/MM | 117 | 0·12 (22/185·6) | 1 | | 1 | |
| Delayed RH5.1/MM | 118 | 0·09 (17/186·5) | 0·24 (-0·44, 0·6) | 0·4011 | 0·29 (-0·43, 0·65) | 0·3353 |
| **Secondary Case Definition** (parasitaemia >**0** parasites/µL) | | | | | | |
| Monthly RH5.1/MM | 117 | 0·23 (39/168·0) | 1 | | 1 | |
| Delayed RH5.1/MM | 118 | 0·15 (26/177·9) | 0·38 (-0·03, 0·62) | 0·0626 | 0·41 (-0·02, 0·65) | 0·0601 |
| **Secondary Case Definition** (parasitaemia >**20,000** parasites/µL) | | | | | | |
| Monthly RH5.1/MM | 117 | 0·08 (16/192·2) | 1 | | 1 | |
| Delayed RH5.1/MM | 118 | 0·05 (10/196·0) | 0·39 (-0·36, 0·72) | 0·2278 | 0·22 (-0·84, 0·67) | 0·5655 |
| **Additional Case Definition** (parasitaemia >**50,000** parasites/µL) | | | | | | |
| Monthly RH5.1/MM | 117 | 0·01 (2/211·0) | 1 | | 1 | |
| Delayed RH5.1/MM | 118 | 0·02 (4/204·9) | -1·04 (-10·14, 0·63) | 0·4100 | -1·82 (-20·61, 0·63) | 0·3176 |
| **Additional Case Definition** (parasitaemia >**100,000** parasites/µL) | | | | | | |
| Monthly RH5.1/MM | 117 | 0·00 (1/212·2) | 1 | | 1 | |
| Delayed RH5.1/MM | 118 | 0·01 (2/207·8) | -1·04 (-21·44, 0·82) | 0·5618 | -1·44 (-45·14, 0·87) | 0·5512 |

#### 3.13 Table S13. Intention-to-treat (ITT) sample endpoint at 6 months: Analysis of vaccine efficacy against all clinical malaria episodes.

A secondary analysis of vaccine efficacy against all clinical malaria episodes was carried out based on the ITT population at the 6-month timepoint, using Cox regression models with a robust standard error to account for multiple episodes. Episodes occurring within 14 days of a previous episode were classed as the same event. For participants without an episode of clinical malaria, their time was censored at the date of their withdrawal or the date of their 6-month blood sampling (noting no deaths have occurred in this trial). Clinical malaria was defined as presence of axillary temperature ≥37·5°C and/or history of fever within the last 24 hours, *P. falciparum* asexual parasitaemia concentration according to case definition. Vaccine Efficacy = 1 – hazard ratio. 95% CI = 95% Confidence Interval. * Adjusted vaccine efficacy: includes covariates of total number of rounds of SMC received, ITN use (adequate or not) the night before screening visit, and age categories 5-8 months, 9-12 months, 13-17 months. Excludes 5 children with missing data on covariates: 1 in the Delayed Rabies arm, 3 in the Delayed RH5.1/MM arm and 1 in the Monthly RH5.1/MM arm. ** Event rate per 100 days (where event is all episodes of clinical malaria) is also reported for information only, calculated as: (the number of events / [child days/100]).

| Groups | N | Number of malaria episodes per child n (%) | | | | Event rate per 100 days ** | VACCINE EFFICACY (95% CI) | p | VACCINE EFFICACY (95% CI) [adjusted*] | p [adjusted] |
|---|---|---|---|---|---|---|---|---|---|---|
| | | 0 | 1 | 2 | 3 | | | | | |
| **Primary Case Definition** (parasitaemia >**5000** parasites/µL) | | | | | | | | | | |
| Rabies controls (combined) | 119 | 85 (71·4) | 27 (22·7) | 7 (5·9) | 0 (0·0) | 0·20 (41/209·3) | 1 | | 1 | |
| Delayed RH5.1/MM | 118 | 101 (85·6) | 16 (13·6) | 1 (0·9) | 0 (0·0) | 0·09 (18/209·7) | 0·56 (0·25, 0·75) | 0·0029 | 0·55 (0·24, 0·74) | 0·0033 |
| Monthly RH5.1/MM | 117 | 95 (81·2) | 20 (17·1) | 2 (1·7) | 0 (0·0) | 0·11 (24/213·8) | 0·42 (0·05, 0·65) | 0·0292 | 0·42 (0·06, 0·64) | 0·0285 |
| **Secondary Case Definition** (parasitaemia >**0** parasites/µL) | | | | | | | | | | |
| Rabies controls (combined) | 119 | 79 (66·4) | 33 (27·7) | 6 (5·0) | 1 (0·8) | 0·23 (48/209·3) | 1 | | 1 | |
| Delayed RH5.1/MM | 118 | 92 (78·0) | 25 (21·2) | 1 (0·9) | 0 (0·0) | 0·13 (27/209·7) | 0·44 (0·13, 0·64) | 0·0102 | 0·41 (0·09, 0·62) | 0·0179 |
| Monthly RH5.1/MM | 117 | 78 (66·7) | 36 (30·8) | 3 (2·6) | 0 (0·0) | 0·20 (42/213·8) | 0·14 (-0·26, 0·42) | 0·4424 | 0·12 (-0·3, 0·41) | 0·5222 |
| **Secondary Case Definition** (parasitaemia >**20,000** parasites/µL) | | | | | | | | | | |
| Rabies controls (combined) | 119 | 91 (76·5) | 22 (18·5) | 6 (5·0) | 0 (0·0) | 0·16 (34/209·3) | 1 | | 1 | |
| Delayed RH5.1/MM | 118 | 108 (91·5) | 10 (8·5) | 0 (0·0) | 0 (0·0) | 0·05 (10/209·7) | 0·71 (0·42, 0·85) | 0·0004 | 0·68 (0·37, 0·84) | 0·0011 |
| Monthly RH5.1/MM | 117 | 101 (86·3) | 14 (12·0) | 2 (1·7) | 0 (0·0) | 0·08 (18/213·8) | 0·48 (0·06, 0·71) | 0·0301 | 0·51 (0·13, 0·72) | 0·0137 |
| **Additional Case Definition** (parasitaemia >**50,000** parasites/µL) | | | | | | | | | | |
| Rabies controls (combined) | 119 | 100 (84·0) | 15 (12·6) | 4 (3·4) | 0 (0·0) | 0·11 (23/209·3) | 1 | | 1 | |
| Delayed RH5.1/MM | 118 | 114 (96·6) | 4 (3·4) | 0 (0·0) | 0 (0·0) | 0·02 (4/209·7) | 0·83 (0·50, 0·94) | 0·0012 | 0·81 (0·42, 0·94) | 0·0035 |
| Monthly RH5.1/MM | 117 | 115 (98·3) | 2 (1·7) | 0 (0·0) | 0 (0·0) | 0·01 (2/213·8) | 0·91 (0·64, 0·98) | 0·0008 | 0·92 (0·66, 0·98) | 0·0006 |
| **Additional Case Definition** (parasitaemia >**100,000** parasites/µL) | | | | | | | | | | |
| Rabies controls (combined) | 119 | 108 (90·8) | 11 (9·2) | 0 (0·0) | 0 (0·0) | 0·05 (11/209·3) | 1 | | 1 | |
| Delayed RH5.1/MM | 118 | 116 (98·3) | 2 (1·7) | 0 (0·0) | 0 (0·0) | 0·01 (2/209·7) | 0·82 (0·20, 0·96) | 0·0242 | 0·80 (0·07, 0·96) | 0·0406 |
| Monthly RH5.1/MM | 117 | 116 (99·2) | 1 (0·9) | 0 (0·0) | 0 (0·0) | 0·00 (1/213·8) | 0·91 (0·32, 0·99) | 0·0199 | 0·91 (0·34, 0·99) | 0·0184 |

##### 3.14 Table S14. Intention-to-treat (ITT) sample endpoint at 6 months: Analysis of vaccine efficacy against all clinical malaria episodes comparing between the two RH5.1/Matrix-M regimens.

A secondary analysis of vaccine efficacy against all clinical malaria episodes comparing between the RH5.1/Matrix-M regimens was carried out based on the ITT population at the 6-month timepoint, using Cox regression models with a robust standard error to account for multiple episodes. Episodes occurring within 14 days of a previous episode were classed as the same event. For participants without an episode of clinical malaria, their time was censored at the date of their withdrawal or the date of their 6-month blood sampling (noting no deaths have occurred in this trial). Clinical malaria was defined as presence of axillary temperature ≥37·5°C and/or history of fever within the last 24 hours, *P. falciparum* asexual parasitaemia concentration according to case definition. Vaccine Efficacy = 1 – hazard ratio. 95% CI = 95% Confidence Interval. * Adjusted vaccine efficacy: includes covariates of total number of rounds of SMC received, ITN use (adequate or not) the night before screening visit, and age categories 5-8 months, 9-12 months, 13-17 months. Excludes 4 children with missing data on covariates: 3 in the Delayed RH5.1/MM arm and 1 in the Monthly RH5.1/MM arm. ** Event rate per 100 days (where event is all episodes of clinical malaria) is also reported for information only, calculated as: (the number of events / [child days/100]).

| Groups | N | Number of malaria episodes per child n (%) | | | | Event rate per 100 days ** | VACCINE EFFICACY (95% CI) | p | VACCINE EFFICACY (95% CI) [adjusted*] | p [adjusted] |
|---|---|---|---|---|---|---|---|---|---|---|
| | | 0 | 1 | 2 | 3 | | | | | |
| **Primary Case Definition** (parasitaemia >**5000** parasites/µL) | | | | | | | | | | |
| Monthly RH5.1/MM | 117 | 95 (81·2) | 20 (17·1) | 2 (1·7) | 0 (0·0) | 0·11 (24/213·8) | 1 | | 1 | |
| Delayed RH5.1/MM | 118 | 101 (85·6) | 16 (13·6) | 1 (0·9) | 0 (0·0) | 0·09 (18/209·7) | 0·24 (-0·38, 0·58) | 0·3672 | 0·3 (-0·37, 0·64) | 0·3000 |
| **Secondary Case Definition** (parasitaemia >**0** parasites/µL) | | | | | | | | | | |
| Monthly RH5.1/MM | 117 | 78 (66·7) | 36 (30·8) | 3 (2·6) | 0 (0·0) | 0·20 (42/213·8) | 1 | | 1 | |
| Delayed RH5.1/MM | 118 | 92 (78·0) | 25 (21·2) | 1 (0·9) | 0 (0·0) | 0·13 (27/209·7) | 0·35 (-0·01, 0·58) | 0·0551 | 0·39 (0·00, 0·63) | 0·0485 |
| **Secondary Case Definition** (parasitaemia >**20,000** parasites/µL) | | | | | | | | | | |
| Monthly RH5.1/MM | 117 | 101 (86·3) | 14 (12·0) | 2 (1·7) | 0 (0·0) | 0·08 (18/213·8) | 1 | | 1 | |
| Delayed RH5.1/MM | 118 | 108 (91·5) | 10 (8·5) | 0 (0·0) | 0 (0·0) | 0·05 (10/209·7) | 0·44 (-0·20, 0·74) | 0·1384 | 0·31 (-0·56, 0·7) | 0·3705 |
| **Additional Case Definition** (parasitaemia >**50,000** parasites/µL) | | | | | | | | | | |
| Monthly RH5.1/MM | 117 | 115 (98·3) | 2 (1·7) | 0 (0·0) | 0 (0·0) | 0·01 (2/213·8) | 1 | | 1 | |
| Delayed RH5.1/MM | 118 | 114 (96·6) | 4 (3·4) | 0 (0·0) | 0 (0·0) | 0·02 (4/209·7) | -1·03 (-9·93, 0·62) | 0·4090 | -1·78 (-19·21, 0·62) | 0·3119 |
| **Additional Case Definition** (parasitaemia >**100,000** parasites/µL) | | | | | | | | | | |
| Monthly RH5.1/MM | 117 | 116 (99·2) | 1 (0·9) | 0 (0·0) | 0 (0·0) | 0·00 (1/213·8) | 1 | | 1 | |
| Delayed RH5.1/MM | 118 | 116 (98·3) | 2 (1·7) | 0 (0·0) | 0 (0·0) | 0·01 (2/209·7) | -1·04 (-21·52, 0·82) | 0·5604 | -1·44 (-42·3, 0·86) | 0·5421 |

#### 3.15 Table S15: Per-protocol sample endpoint at 3 months – Analysis of time to first episode of clinical malaria from 14 days to 3 months post-third vaccination.

Cox regression models were used to analyse the secondary endpoint of time to first episode of clinical malaria based on the per-protocol population at the 3-month analysis timepoint. Clinical malaria is defined as measured axillary temperature ≥37·5°C and/or reported history of fever within the last 24 hours, and *P. falciparum* asexual parasitaemia concentration according to each case definition. Vaccine Efficacy = 1 – hazard ratio. 95% CI = 95% Confidence Interval. * Adjusted vaccine efficacy: includes covariates of total number of rounds of SMC received, ITN use (adequate or not) the night before the screening visit, and age categories 5-8 months, 9-12 months, 13-17 months. Excludes 5 children with missing data on covariates: 1 in the Delayed Rabies arm, 3 in the Delayed RH5.1/Matrix-M arm and 1 in the Monthly RH5.1/Matrix-M arm. ** Event rate per 100 days (where event is the first episode of clinical malaria) is also reported for information only, calculated as: (the number of events / [child days/100]).

| **Groups** | **N** | **Event rate per 100 days **** | **VACCINE EFFICACY (95% CI)** | **p** | **VACCINE EFFICACY (95% CI) [adjusted*]** | **p [adjusted]** |
|---|---|---|---|---|---|---|
| **Primary Case Definition** (parasitaemia >**5000** parasites/µL) | | | | | | |
| Rabies controls (combined) | 112 | 0·38 (32/83·9) | 1 | | 1 | |
| Delayed RH5.1/MM | 114 | 0·17 (16/96·4) | 0·56 (0·21, 0·76) | 0·0062 | 0·57 (0·21, 0·77) | 0·0070 |
| Monthly RH5.1/MM | 112 | 0·18 (17/92·9) | 0·52 (0·13, 0·73) | 0·0153 | 0·54 (0·18, 0·74) | 0·0091 |
| **Secondary Case Definition** (parasitaemia >**0** parasites/µL) | | | | | | |
| Rabies controls (combined) | 112 | 0·46 (37/80·3) | 1 | | 1 | |
| Delayed RH5.1/MM | 114 | 0·22 (21/94·4) | 0·52 (0·19, 0·72) | 0·0064 | 0·52 (0·17, 0·72) | 0·0086 |
| Monthly RH5.1/MM | 112 | 0·31 (27/86·7) | 0·32 (-0·12, 0·59) | 0·1260 | 0·33 (-0·12, 0·59) | 0·1252 |
| **Secondary Case Definition** (parasitaemia >**20,000** parasites/µL) | | | | | | |
| Rabies controls (combined) | 112 | 0·31 (27/87·3) | 1 | | 1 | |
| Delayed RH5.1/MM | 114 | 0·09 (9/99·5) | 0·70 (0·37, 0·86) | 0·0015 | 0·69 (0·32, 0·86) | 0·0037 |
| Monthly RH5.1/MM | 112 | 0·15 (14/94·8) | 0·52 (0·09, 0·75) | 0·0254 | 0·56 (0·18, 0·77) | 0·0107 |
| **Additional Case Definition** (parasitaemia >**50,000** parasites/µL) | | | | | | |
| Rabies controls (combined) | 112 | 0·19 (18/93·5) | 1 | | 1 | |
| Delayed RH5.1/MM | 114 | 0·03 (3/102·9) | 0·85 (0·49, 0·96) | 0·0024 | 0·83 (0·39, 0·96) | 0·0068 |
| Monthly RH5.1/MM | 112 | 0·02 (2/101·4) | 0·90 (0·56, 0·98) | 0·0023 | 0·91 (0·59, 0·98) | 0·0016 |
| **Additional Case Definition** (parasitaemia >**100,000** parasites/µL) | | | | | | |
| Rabies controls (combined) | 112 | 0·10 (10/97·6) | 1 | | 1 | |
| Delayed RH5.1/MM | 114 | 0·01 (1/103·9) | 0·91 (0·27, 0·99) | 0·0237 | 0·90 (0·17, 0·99) | 0·0331 |
| Monthly RH5.1/MM | 112 | 0·01 (1/101·7) | 0·90 (0·25, 0·99) | 0·0259 | 0·91 (0·29, 0·99) | 0·0230 |

#### 3.16 Figure S1: Kaplan-Meier curves showing time to first episode of clinical malaria from 14 days to 3 months post-third vaccination.

Analysis was based on the per-protocol population. (**A**) Primary case definition of clinical malaria with parasitaemia >5000 parasites/µL. (**B**) Secondary case definition of clinical malaria with parasitaemia >0 parasites/µL, or (**C**) >20,000 parasites/µL. (**D**) Additional case definition of clinical malaria with parasitaemia >50,000 parasites/µL, or (**E**) >100,000 parasites/µL.

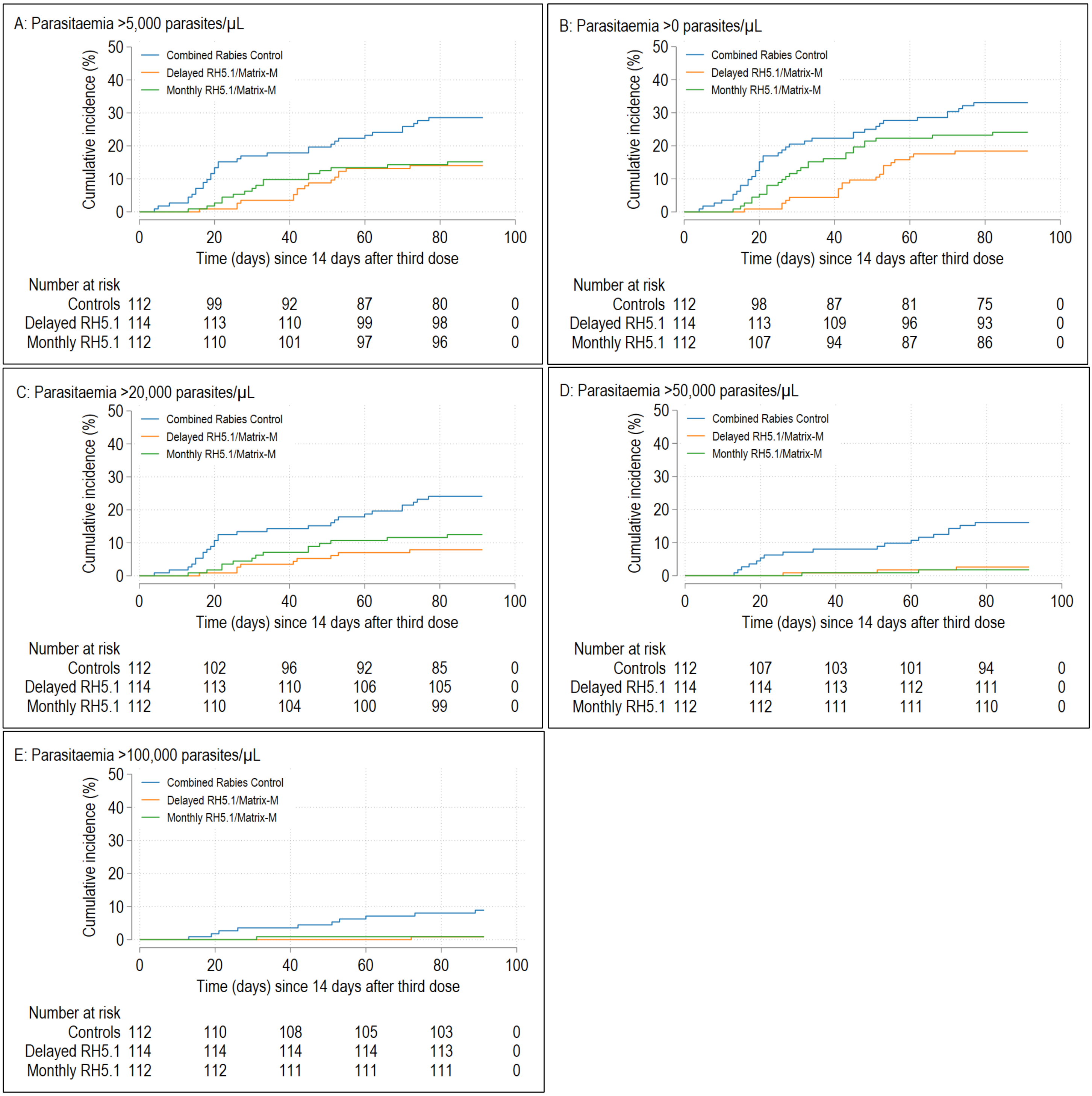

#### 3.17 Table S16: Intention-to-treat (ITT) sample endpoint at 3 months – Analysis of time to first episode of clinical malaria from 14 days to 3 months post-third vaccination.

Cox regression models were used to analyse the secondary endpoint of time to first episode of clinical malaria based on the ITT population at the 3-month analysis timepoint. Clinical malaria is defined as measured axillary temperature ≥37·5°C and/or reported history of fever within the last 24 hours, and *P. falciparum* asexual parasitaemia concentration according to each case definition. Vaccine Efficacy = 1 – hazard ratio. 95% CI = 95% Confidence Interval. * Adjusted vaccine efficacy: includes covariates of total number of rounds of SMC received, ITN use (adequate or not) the night before the screening visit, and age categories 5-8 months, 9-12 months, 13-17 months. Excludes 5 children with missing data on covariates: 1 in the Delayed Rabies arm, 3 in the Delayed RH5.1/Matrix-M arm and 1 in the Monthly RH5.1/Matrix-M arm. ** Event rate per 100 days (where event is the first episode of clinical malaria) is also reported for information only, calculated as: (the number of events / [child days/100]).

| Groups | N | Event rate per 100 days ** | VACCINE EFFICACY (95% CI) | p | VACCINE EFFICACY (95% CI) [adjusted*] | p [adjusted] |
|---|---|---|---|---|---|---|
| **Primary Case Definition** (parasitaemia >**5000** parasites/µL) | | | | | | |
| Rabies controls (combined) | 119 | 0·38 (33/85·9) | 1 | | 1 | |
| Delayed RH5.1/MM | 118 | 0·16 (16/97·9) | 0·57 (0·23, 0·76) | 0·0046 | 0·56 (0·19, 0·76) | 0·0088 |
| Monthly RH5.1/MM | 117 | 0·17 (17/97·5) | 0·54 (0·18, 0·74) | 0·0089 | 0·55 (0·19, 0·75) | 0·0075 |
| **Secondary Case Definition** (parasitaemia >**0** parasites/µL) | | | | | | |
| Rabies controls (combined) | 119 | 0·46 (38/82·4) | 1 | | 1 | |
| Delayed RH5.1/MM | 118 | 0·22 (21/95·8) | 0·53 (0·2, 0·72) | 0·0049 | 0·50 (0·15, 0·71) | 0·0111 |
| Monthly RH5.1/MM | 117 | 0·30 (27/91·3) | 0·36 (-0·06, 0·61) | 0·0820 | 0·35 (-0·09, 0·61) | 0·1003 |
| **Secondary Case Definition** (parasitaemia >**20,000** parasites/µL) | | | | | | |
| Rabies controls (combined) | 119 | 0·30 (27/90·1) | 1 | | 1 | |
| Delayed RH5.1/MM | 118 | 0·09 (9/100·9) | 0·70 (0·36, 0·86) | 0·0017 | 0·69 (0·30, 0·86) | 0·0043 |
| Monthly RH5.1/MM | 117 | 0·14 (14/99·4) | 0·53 (0·1, 0·75) | 0·0227 | 0·57 (0·19, 0·77) | 0·0095 |
| **Additional Case Definition** (parasitaemia >**50,000** parasites/µL) | | | | | | |
| Rabies controls (combined) | 119 | 0·19 (18/96·3) | 1 | | 1 | |
| Delayed RH5.1/MM | 118 | 0·03 (3/104·4) | 0·85 (0·48, 0·95) | 0·0026 | 0·83 (0·38, 0·95) | 0·0070 |
| Monthly RH5.1/MM | 117 | 0·02 (2/106·0) | 0·90 (0·56, 0·98) | 0·0021 | 0·91 (0·6, 0·98) | 0·0015 |
| **Additional Case Definition** (parasitaemia >**100,000** parasites/µL) | | | | | | |
| Rabies controls (combined) | 119 | 0·10 (10/100·5) | 1 | | 1 | |
| Delayed RH5.1/MM | 118 | 0·01 (1/105·4) | 0·90 (0·26, 0·99) | 0·0245 | 0·90 (0·16, 0·99) | 0·0341 |
| Monthly RH5.1/MM | 117 | 0·01 (1/106·3) | 0·91 (0·26, 0·99) | 0·0249 | 0·91 (0·29, 0·99) | 0·0222 |

#### 3.18 Table S17: Solicited Adverse Events in the 7 days following first three vaccinations, including day of vaccination.

Definitions of grade severity are provided in **Table S2**. In the Delayed Rabies group (Rabivax-S) 62, 61, and 55 children received doses 1, 2, and 3 respectively; in the Delayed RH5.1/MM group (10µg RH5.1 / 50µg Matrix-M) 119, 118, and 114 received doses 1, 2, and 3 respectively; in the Monthly Rabies group (Rabivax-S) 60, 60, and 57 received 1, 2, and 3 respectively; and in the Monthly RH5.1/MM group (10µg RH5.1 / 50µg Matrix-M) the numbers per group were 120, 119, and 113 who received doses 1, 2, and 3 respectively. Each event is counted at its highest grade over the 7 day follow up for each child.

| Adverse Event | Dose Number | No. of participants in Delayed Rabies control (%) | | | No. of participants in Delayed RH5.1/MM (%) | | | No. of participants in Monthly Rabies control (%) | | | No. of participants in Monthly RH5.1/MM (%) | | |
|---|---|---|---|---|---|---|---|---|---|---|---|---|---|
| | | Mild | Moderate | Severe | Mild | Moderate | Severe | Mild | Moderate | Severe | Mild | Moderate | Severe |
| Pain | 1 | 2 (3·2) | 0 (0·0) | 1 (1·6) | 5 (4·2) | 0 (0·0) | 0 (0·0) | 0 (0·0) | 0 (0·0) | 1 (1·7) | 0 (0·0) | 0 (0·0) | 0 (0·0) |
| | 2 | 0 (0·0) | 0 (0·0) | 1 (1·6) | 5 (4·2) | 1 (0·8) | 0 (0·0) | 0 (0·0) | 0 (0·0) | 0 (0·0) | 1 (0·8) | 0 (0·0) | 0 (0·0) |
| | 3 | 0 (0·0) | 0 (0·0) | 0 (0·0) | 1 (0·9) | 0 (0·0) | 2 (1·8) | 0 (0·0) | 0 (0·0) | 1 (1·8) | 1 (0·9) | 0 (0·0) | 2 (1·8) |
| Swelling | 1 | 1 (1·6) | 0 (0·0) | 0 (0·0) | 0 (0·0) | 0 (0·0) | 0 (0·0) | 0 (0·0) | 0 (0·0) | 0 (0·0) | 0 (0·0) | 0 (0·0) | 0 (0·0) |
| | 2 | 0 (0·0) | 0 (0·0) | 0 (0·0) | 13 (11) | 4 (3·4) | 0 (0·0) | 0 (0·0) | 1 (1·7) | 0 (0·0) | 4 (3·4) | 1 (0·8) | 0 (0·0) |
| | 3 | 0 (0·0) | 0 (0·0) | 0 (0·0) | 0 (0·0) | 2 (1·8) | 0 (0·0) | 0 (0·0) | 0 (0·0) | 0 (0·0) | 0 (0·0) | 0 (0·0) | 0 (0·0) |
| Redness | 1 | 0 (0·0) | 0 (0·0) | 0 (0·0) | 0 (0·0) | 0 (0·0) | 0 (0·0) | 0 (0·0) | 1 (1·7) | 0 (0·0) | 0 (0·0) | 0 (0·0) | 0 (0·0) |
| | 2 | 0 (0·0) | 0 (0·0) | 0 (0·0) | 1 (0·8) | 0 (0·0) | 0 (0·0) | 0 (0·0) | 0 (0·0) | 0 (0·0) | 0 (0·0) | 2 (1·7) | 0 (0·0) |
| | 3 | 0 (0·0) | 0 (0·0) | 0 (0·0) | 0 (0·0) | 0 (0·0) | 0 (0·0) | 0 (0·0) | 0 (0·0) | 0 (0·0) | 0 (0·0) | 0 (0·0) | 0 (0·0) |
| Warmth | 1 | 1 (1·6) | 1 (1·6) | 0 (0·0) | 3 (2·5) | 0 (0·0) | 0 (0·0) | 0 (0·0) | 0 (0·0) | 0 (0·0) | 0 (0·0) | 0 (0·0) | 0 (0·0) |
| | 2 | 0 (0·0) | 0 (0·0) | 0 (0·0) | 2 (1·7) | 0 (0·0) | 0 (0·0) | 0 (0·0) | 0 (0·0) | 0 (0·0) | 4 (3·4) | 0 (0·0) | 0 (0·0) |
| | 3 | 0 (0·0) | 0 (0·0) | 1 (1·8) | 0 (0·0) | 1 (0·9) | 0 (0·0) | 0 (0·0) | 0 (0·0) | 0 (0·0) | 1 (0·9) | 0 (0·0) | 0 (0·0) |
| Fever | 1 | 0 (0·0) | 1 (1·6) | 0 (0·0) | 2 (1·7) | 4 (3·4) | 0 (0·0) | 0 (0·0) | 0 (0·0) | 0 (0·0) | 3 (2·5) | 1 (0·8) | 0 (0·0) |
| | 2 | 2 (3·3) | 0 (0·0) | 0 (0·0) | 15 (12·7) | 11 (9·3) | 1 (0·8) | 0 (0·0) | 1 (1·7) | 0 (0·0) | 15 (12·6) | 9 (7·6) | 2 (1·7) |
| | 3 | 1 (1·8) | 0 (0·0) | 0 (0·0) | 14 (12·3) | 7 (6·1) | 0 (0·0) | 0 (0·0) | 0 (0·0) | 0 (0·0) | 4 (3·5) | 9 (8) | 0 (0·0) |
| Irritable | 1 | 0 (0·0) | 0 (0·0) | 0 (0·0) | 0 (0·0) | 0 (0·0) | 0 (0·0) | 0 (0·0 | 0 (0·0) | 0 (0·0) | 0 (0·0) | 0 (0·0) | 0 (0·0) |
| | 2 | 1 (1·6) | 0 (0·0) | 0 (0·0) | 2 (1·7) | 0 (0·0) | 0 (0·0) | 1 (1·7) | 0 (0·0) | 0 (0·0) | 0 (0·0) | 0 (0·0) | 0 (0·0) |
| | 3 | 1 (1·8) | 0 (0·0) | 0 (0·0) | 0 (0·0) | 1 (0·9) | 0 (0·0) | 1 (1·8) | 0 (0·0) | 0 (0·0) | 1 (0·9) | 0 (0·0) | 0 (0·0) |

| | | | | | | | | | | | | | |
|---|---|---|---|---|---|---|---|---|---|---|---|---|---|
| Drowsiness | 1 | 0 (0·0) | 0 (0·0) | 0 (0·0) | 4 (3·4) | 0 (0·0) | 0 (0·0) | 1 (1·7) | 0 (0·0) | 0 (0·0) | 0 (0·0) | 0 (0·0) | 0 (0·0) |
| | 2 | 0 (0·0) | 0 (0·0) | 0 (0·0) | 2 (1·7) | 1 (0·8) | 0 (0·0) | 0 (0·0 | 0 (0·0) | 0 (0·0) | 1 (0·8) | 0 (0·0) | 0 (0·0) |
| | 3 | 0 (0·0) | 0 (0·0) | 0 (0·0) | 0 (0·0) | 0 (0·0) | 0 (0·0) | 0 (0·0) | 0 (0·0) | 0 (0·0) | 1 (0·9) | 0 (0·0) | 0 (0·0) |
| Loss of appetite | 1 | 0 (0·0) | 1 (1·6) | 0 (0·0) | 2 (1·7) | 0 (0·0) | 0 (0·0) | 0 (0·0) | 0 (0·0) | 0 (0·0) | 0 (0·0) | 0 (0·0) | 0 (0·0) |
| | 2 | 2 (3·3) | 0 (0·0) | 0 (0·0) | 3 (2·5) | 0 (0·0) | 1 (0·8) | 0 (0·0 | 1 (1·7) | 0 (0·0) | 1 (0·8) | 0 (0·0) | 0 (0·0) |
| | 3 | 1 (1·8) | 0 (0·0) | 0 (0·0) | 0 (0·0) | 1 (0·9) | 0 (0·0) | 0 (0·0 | 0 (0·0) | 0 (0·0) | 1 (0·9) | 0 (0·0) | 0 (0·0) |

#### 3.19 Table S18: Unsolicited Adverse Events within 28 days of vaccine by dose and group.

Number and percentage of children with at least one AE of each type per dose and arm. Includes all children who received at least one dose of vaccine, by dose. All unsolicited AEs were classified as unrelated to vaccination, with the exception of one episode of moderate fever in the delayed third-dose group (occurring within 28 days of vaccination).

| Arm | Delayed Rabies Control | | | Delayed RH5.1/Matrix-M | | | Monthly Rabies Control | | | Monthly RH5.1/Matrix-M | | |
|---|---|---|---|---|---|---|---|---|---|---|---|---|
| **Dose** | **1** | **2** | **3** | **1** | **2** | **3** | **1** | **2** | **3** | **1** | **2** | **3** |
| **Number of children followed** | **62** | **61** | **55** | **119** | **118** | **114** | **60** | **60** | **57** | **120** | **119** | **113** |
| Abdominal pain | 0 (0·0) | 0 (0·0) | 0 (0·0) | 1 (0·8) | 0 (0·0) | 0 (0·0) | 0 (0·0) | 0 (0·0) | 0 (0·0) | 0 (0·0) | 0 (0·0) | 0 (0·0) |
| Anaemia | 6 (9·7) | 11 (18) | 6 (10·9) | 20 (16·8) | 12 (10·2) | 10 (8·8) | 3 (5) | 14 (23·3) | 1 (1·8) | 5 (4·2) | 27 (22·7) | 0 (0·0) |
| Anal candidiasis | 0 (0·0) | 0 (0·0) | 0 (0·0) | 0 (0·0) | 0 (0·0) | 0 (0·0) | 0 (0·0) | 0 (0·0) | 1 (1·8) | 0 (0·0) | 1 (0·8) | 0 (0·0) |
| Arthropod sting | 0 (0·0) | 0 (0·0) | 0 (0·0) | 1 (0·8) | 0 (0·0) | 0 (0·0) | 0 (0·0) | 0 (0·0) | 0 (0·0) | 0 (0·0) | 0 (0·0) | 0 (0·0) |
| Bronchiolitis | 0 (0·0) | 1 (1·6) | 0 (0·0) | 0 (0·0) | 0 (0·0) | 0 (0·0) | 0 (0·0) | 0 (0·0) | 0 (0·0) | 0 (0·0) | 0 (0·0) | 0 (0·0) |
| Bronchitis | 18 (29) | 16 (26·2) | 20 (36·4) | 49 (41·2) | 32 (27·1) | 32 (28·1) | 14 (23·3) | 17 (28·3) | 21 (36·8) | 13 (10·8) | 35 (29·4) | 46 (40·7) |
| Candida infection | 0 (0·0) | 0 (0·0) | 0 (0·0) | 0 (0·0) | 0 (0·0) | 0 (0·0) | 0 (0·0) | 0 (0·0) | 0 (0·0) | 1 (0·8) | 0 (0·0) | 0 (0·0) |
| Cellulitis | 0 (0·0) | 0 (0·0) | 0 (0·0) | 0 (0·0) | 0 (0·0) | 1 (0·9) | 0 (0·0) | 0 (0·0) | 0 (0·0) | 0 (0·0) | 0 (0·0) | 0 (0·0) |
| Conjunctivitis | 7 (11·3) | 3 (4·9) | 0 (0·0) | 6 (5) | 3 (2·5) | 0 (0·0) | 2 (3·3) | 0 (0·0) | 0 (0·0) | 3 (2·5) | 3 (2·5) | 4 (3·5) |
| Conjunctivitis – bacterial | 0 (0·0) | 1 (1·6) | 0 (0·0) | 0 (0·0) | 0 (0·0) | 0 (0·0) | 0 (0·0) | 0 (0·0) | 0 (0·0) | 0 (0·0) | 0 (0·0) | 0 (0·0) |
| Dermatitis | 0 (0·0) | 0 (0·0) | 0 (0·0) | 0 (0·0) | 1 (0·8) | 0 (0·0) | 0 (0·0) | 0 (0·0) | 0 (0·0) | 1 (0·8) | 2 (1·7) | 0 (0·0) |
| Dermatitis atopic | 0 (0·0) | 0 (0·0) | 1 (1·8) | 0 (0·0) | 1 (0·8) | 0 (0·0) | 0 (0·0) | 0 (0·0) | 0 (0·0) | 0 (0·0) | 0 (0·0) | 0 (0·0) |
| Dermatitis diaper | 0 (0·0) | 0 (0·0) | 0 (0·0) | 0 (0·0) | 0 (0·0) | 0 (0·0) | 0 (0·0) | 0 (0·0) | 0 (0·0) | 1 (0·8) | 0 (0·0) | 0 (0·0) |
| Ear infection | 0 (0·0) | 0 (0·0) | 0 (0·0) | 0 (0·0) | 0 (0·0) | 0 (0·0) | 1 (1·7) | 0 (0·0) | 0 (0·0) | 0 (0·0) | 2 (1·7) | 0 (0·0) |
| Ear injury | 0 (0·0) | 0 (0·0) | 1 (1·8) | 0 (0·0) | 0 (0·0) | 0 (0·0) | 1 (1·7) | 0 (0·0) | 0 (0·0) | 0 (0·0) | 0 (0·0) | 0 (0·0) |
| Enteritis | 9 (14·5) | 19 (31·1) | 4 (7·3) | 24 (20·2) | 26 (22) | 20 (17·5) | 20 (33·3) | 21 (35) | 9 (15·8) | 31 (25·8) | 42 (35·3) | 24 (21·2) |
| Furuncle | 1 (1·6) | 2 (3·3) | 0 (0·0) | 1 (0·8) | 5 (4·2) | 2 (1·8) | 2 (3·3) | 0 (0·0) | 0 (0·0) | 6 (5) | 3 (2·5) | 0 (0·0) |
| Gastroenteritis | 0 (0·0) | 0 (0·0) | 0 (0·0) | 0 (0·0) | 0 (0·0) | 0 (0·0) | 1 (1·7) | 0 (0·0) | 0 (0·0) | 0 (0·0) | 1 (0·8) | 1 (0·9) |
| Gastrointestinal - candidiasis | 0 (0·0) | 1 (1·6) | 0 (0·0) | 0 (0·0) | 5 (4·2) | 0 (0·0) | 2 (3·3) | 1 (1·7) | 2 (3·5) | 1 (0·8) | 4 (3·4) | 0 (0·0) |
| Impetigo | 0 (0·0) | 0 (0·0) | 0 (0·0) | 0 (0·0) | 0 (0·0) | 1 (0·9) | 0 (0·0) | 0 (0·0) | 1 (1·8) | 0 (0·0) | 0 (0·0) | 1 (0·9) |
| Infection parasitic | 1 (1·6) | 1 (1·6) | 0 (0·0) | 1 (0·8) | 1 (0·8) | 0 (0·0) | 1 (1·7) | 0 (0·0) | 2 (3·5) | 1 (0·8) | 5 (4·2) | 0 (0·0) |
| Laryngitis | 0 (0·0) | 0 (0·0) | 0 (0·0) | 0 (0·0) | 1 (0·8) | 0 (0·0) | 0 (0·0) | 0 (0·0) | 0 (0·0) | 0 (0·0) | 0 (0·0) | 1 (0·9) |
| Malaria | 1 (1·6) | 0 (0·0) | 5 (9·1) | 4 (3·4) | 1 (0·8) | 6 (5·3) | 2 (3·3) | 5 (8·3) | 5 (8·8) | 5 (4·2) | 17 (14·3) | 4 (3·5) |
| Myositis | 0 (0·0) | 1 (1·6) | 0 (0·0) | 0 (0·0) | 0 (0·0) | 0 (0·0) | 0 (0·0) | 0 (0·0) | 0 (0·0) | 0 (0·0) | 0 (0·0) | 0 (0·0) |
| Oral candidiasis | 0 (0·0) | 0 (0·0) | 0 (0·0) | 0 (0·0) | 0 (0·0) | 1 (0·9) | 0 (0·0) | 1 (1·7) | 0 (0·0) | 0 (0·0) | 2 (1·7) | 0 (0·0) |
| Otitis media acute | 1 (1·6) | 4 (6·6) | 4 (7·3) | 4 (3·4) | 8 (6·8) | 4 (3·5) | 0 (0·0) | 4 (6·7) | 5 (8·8) | 8 (6·7) | 12 (10·1) | 5 (4·4) |
| Parasitic gastroenteritis | 0 (0·0) | 0 (0·0) | 0 (0·0) | 0 (0·0) | 1 (0·8) | 0 (0·0) | 0 (0·0) | 0 (0·0) | 0 (0·0) | 0 (0·0) | 2 (1·7) | 1 (0·9) |
| Parotitis | 0 (0·0) | 0 (0·0) | 0 (0·0) | 1 (0·8) | 0 (0·0) | 0 (0·0) | 0 (0·0) | 0 (0·0) | 0 (0·0) | 0 (0·0) | 0 (0·0) | 0 (0·0) |
| Pharyngitis | 5 (8·1) | 2 (3·3) | 0 (0·0) | 2 (1·7) | 1 (0·8) | 2 (1·8) | 1 (1·7) | 0 (0·0) | 0 (0·0) | 4 (3·3) | 3 (2·5) | 2 (1·8) |
| Poisoning | 0 (0·0) | 0 (0·0) | 0 (0·0) | 0 (0·0) | 0 (0·0) | 0 (0·0) | 0 (0·0) | 0 (0·0) | 0 (0·0) | 0 (0·0) | 0 (0·0) | 1 (0·9) |
| Prurigo | 0 (0·0) | 0 (0·0) | 0 (0·0) | 0 (0·0) | 0 (0·0) | 0 (0·0) | 1 (1·7) | 1 (1·7) | 3 (5·3) | 6 (5) | 6 (5) | 1 (0·9) |

| **Arm** | **Delayed Rabies Control** | | | **Delayed RH5.1/Matrix-M** | | | **Monthly Rabies Control** | | | **Monthly RH5.1/Matrix-M** | | |
|---|---|---|---|---|---|---|---|---|---|---|---|---|
| **Dose** | **1** | **2** | **3** | **1** | **2** | **3** | **1** | **2** | **3** | **1** | **2** | **3** |
| **Number of children followed** | **62** | **61** | **55** | **119** | **118** | **114** | **60** | **60** | **57** | **120** | **119** | **113** |
| Pyoderma | 1 (1·6) | 3 (4·9) | 2 (3·6) | 2 (1·7) | 8 (6·8) | 11 (9·6) | 9 (15) | 4 (6·7) | 0 (0·0) | 19 (15·8) | 10 (8·4) | 8 (7·1) |
| Pyrexia | 0 (0·0) | 0 (0·0) | 0 (0·0) | 1 (0·8) | 0 (0·0) | 0 (0·0) | 0 (0·0) | 0 (0·0) | 0 (0·0) | 0 (0·0) | 0 (0·0) | 0 (0·0) |
| Rhinitis | 18(29) | 11(18) | 4 (7·3) | 34 (28·6) | 25 (21·2) | 11 (9·6) | 6 (10) | 3 (5) | 1 (1·8) | 26 (21·7) | 8 (6·7) | 10 (8·8) |
| Skin candida | 0 (0·0) | 0 (0·0) | 0 (0·0) | 0 (0·0) | 0 (0·0) | 0 (0·0) | 1 (1·7) | 0 (0·0) | 0 (0·0) | 1 (0·8) | 0 (0·0) | 0 (0·0) |
| Staphylococcal skin infection | 0 (0·0) | 1 (1·6) | 0 (0·0) | 0 (0·0) | 0 (0·0) | 0 (0·0) | 0 (0·0) | 0 (0·0) | 0 (0·0) | 0 (0·0) | 0 (0·0) | 0 (0·0) |
| Thermal burn | 0 (0·0) | 0 (0·0) | 0 (0·0) | 0 (0·0) | 0 (0·0) | 0 (0·0) | 0 (0·0) | 0 (0·0) | 0 (0·0) | 0 (0·0) | 1 (0·8) | 1 (0·9) |
| Urinary tract infection | 0 (0·0) | 0 (0·0) | 0 (0·0) | 0 (0·0) | 1 (0·8) | 0 (0·0) | 0 (0·0) | 0 (0·0) | 0 (0·0) | 0 (0·0) | 0 (0·0) | 0 (0·0) |
| Wound | 0 (0·0) | 1 (1·6) | 0 (0·0) | 0 (0·0) | 0 (0·0) | 0 (0·0) | 1 (1·7) | 0 (0·0) | 0 (0·0) | 1 (0·8) | 0 (0·0) | 0 (0·0) |

#### 3.20 Table S19: Unsolicited Adverse Events within 28 days of vaccine doses 1-3 by group.

Unsolicited AEs by severity of event combined for all three vaccine doses. Data show the number and percentage of children with at least one AE of each type per severity level and arm. Includes all children in the intention-to-treat sample. Mod. = Moderate; Sev. = Severe.

| **Arm** | **Delayed Rabies Control** | | | **Delayed RH5.1/Matrix-M** | | | **Monthly Rabies Control** | | | **Monthly RH5.1/Matrix-M** | | |
|---|---|---|---|---|---|---|---|---|---|---|---|---|
| **Grade** | Mild | Mod. | Sev. | Mild | Mod. | Sev. | Mild | Mod. | Sev. | Mild | Mod. | Sev. |
| **Number of children followed** | **62** | | | **119** | | | **60** | | | **120** | | |
| Abdominal pain | 0 (0·0) | 0 (0·0) | 0 (0·0) | 0 (0·0) | 1 (0·8) | 0 (0·0) | 0 (0·0) | 0 (0·0) | 0 (0·0) | 0 (0·0) | 0 (0·0) | 0 (0·0) |
| Anaemia | 7 (11·3) | 14 (22·6) | 2 (3·2) | 9 (7·6) | 31 (26·1) | 2 (1·7) | 3 (5) | 15 (25) | 0 (0·0) | 2 (1·7) | 28 (23·3) | 2 (1·7) |
| Anal candidiasis | 0 (0·0) | 0 (0·0) | 0 (0·0) | 0 (0·0) | 0 (0·0) | 0 (0·0) | 0 (0·0) | 1 (1·7) | 0 (0·0) | 0 (0·0) | 1 (0·8) | 0 (0·0) |
| Arthropod sting | 0 (0·0) | 0 (0·0) | 0 (0·0) | 0 (0·0) | 1 (0·8) | 0 (0·0) | 0 (0·0) | 0 (0·0) | 0 (0·0) | 0 (0·0) | 0 (0·0) | 0 (0·0) |
| Bronchiolitis | 0 (0·0) | 1 (1·6) | 0 (0·0) | 0 (0·0) | 0 (0·0) | 0 (0·0) | 0 (0·0) | 0 (0·0) | 0 (0·0) | 0 (0·0) | 0 (0·0) | 0 (0·0) |
| Bronchitis | 0 (0·0) | 41 (66·1) | 0 (0·0) | 2 (1·7) | 87 (73·1) | 0 (0·0) | 0 (0·0) | 38 (63·3) | 0 (0·0) | 0 (0·0) | 74 (61·7) | 0 (0·0) |
| Candida infection | 0 (0·0) | 0 (0·0) | 0 (0·0) | 0 (0·0) | 0 (0·0) | 0 (0·0) | 0 (0·0) | 0 (0·0) | 0 (0·0) | 0 (0·0) | 1 (0·8) | 0 (0·0) |
| Cellulitis | 0 (0·0) | 0 (0·0) | 0 (0·0) | 0 (0·0) | 1 (0·8) | 0 (0·0) | 0 (0·0) | 0 (0·0) | 0 (0·0) | 0 (0·0) | 0 (0·0) | 0 (0·0) |
| Conjunctivitis | 0 (0·0) | 10 (16·1) | 0 (0·0) | 0 (0·0) | 9 (7·6) | 0 (0·0) | 0 (0·0) | 2 (3·3) | 0 (0·0) | 0 (0·0) | 9 (7·5) | 0 (0·0) |
| Conjunctivitis - bacterial | 0 (0·0) | 1 (1·6) | 0 (0·0) | 0 (0·0) | 0 (0·0) | 0 (0·0) | 0 (0·0) | 0 (0·0) | 0 (0·0) | 0 (0·0) | 0 (0·0) | 0 (0·0) |
| Dermatitis | 0 (0·0) | 0 (0·0) | 0 (0·0) | 0 (0·0) | 1 (0·8) | 0 (0·0) | 0 (0·0) | 0 (0·0) | 0 (0·0) | 0 (0·0) | 3 (2·5) | 0 (0·0) |
| Dermatitis atopic | 0 (0·0) | 1 (1·6) | 0 (0·0) | 0 (0·0) | 1 (0·8) | 0 (0·0) | 0 (0·0) | 0 (0·0) | 0 (0·0) | 0 (0·0) | 0 (0·0) | 0 (0·0) |
| Dermatitis diaper | 0 (0·0) | 0 (0·0) | 0 (0·0) | 0 (0·0) | 0 (0·0) | 0 (0·0) | 0 (0·0) | 0 (0·0) | 0 (0·0) | 0 (0·0) | 1 (0·8) | 0 (0·0) |
| Ear infection | 0 (0·0) | 0 (0·0) | 0 (0·0) | 0 (0·0) | 0 (0·0) | 0 (0·0) | 0 (0·0) | 1 (1·7) | 0 (0·0) | 0 (0·0) | 2 (1·7) | 0 (0·0) |
| Ear injury | 0 (0·0) | 1 (1·6) | 0 (0·0) | 0 (0·0) | 0 (0·0) | 0 (0·0) | 0 (0·0) | 1 (1·7) | 0 (0·0) | 0 (0·0) | 0 (0·0) | 0 (0·0) |
| Enteritis | 2 (3·2) | 25 (40·3) | 0 (0·0) | 0 (0·0) | 54 (45·4) | 0 (0·0) | 0 (0·0) | 36 (60) | 0 (0·0) | 0 (0·0) | 72 (60) | 0 (0·0) |
| Furuncle | 0 (0·0) | 3 (4·8) | 0 (0·0) | 0 (0·0) | 8 (6·7) | 0 (0·0) | 0 (0·0) | 2 (3·3) | 0 (0·0) | 0 (0·0) | 9 (7·5) | 0 (0·0) |
| Gastroenteritis | 0 (0·0) | 0 (0·0) | 0 (0·0) | 0 (0·0) | 0 (0·0) | 0 (0·0) | 0 (0·0) | 1 (1·7) | 0 (0·0) | 0 (0·0) | 2 (1·7) | 0 (0·0) |
| Gastrointestinal - candidiasis | 0 (0·0) | 1 (1·6) | 0 (0·0) | 0 (0·0) | 5 (4·2) | 0 (0·0) | 0 (0·0) | 5 (8·3) | 0 (0·0) | 0 (0·0) | 5 (4·2) | 0 (0·0) |
| Impetigo | 0 (0·0) | 0 (0·0) | 0 (0·0) | 0 (0·0) | 1 (0·8) | 0 (0·0) | 0 (0·0) | 1 (1·7) | 0 (0·0) | 0 (0·0) | 1 (0·8) | 0 (0·0) |
| Infection parasitic | 0 (0·0) | 2 (3·2) | 0 (0·0) | 0 (0·0) | 2 (1·7) | 0 (0·0) | 0 (0·0) | 3 (5) | 0 (0·0) | 0 (0·0) | 6 (5) | 0 (0·0) |
| Laryngitis | 0 (0·0) | 0 (0·0) | 0 (0·0) | 0 (0·0) | 1 (0·8) | 0 (0·0) | 0 (0·0) | 0 (0·0) | 0 (0·0) | 0 (0·0) | 1 (0·8) | 0 (0·0) |
| Malaria | 0 (0·0) | 6 (9·7) | 0 (0·0) | 0 (0·0) | 10 (8·4) | 0 (0·0) | 0 (0·0) | 11 (18·3) | 0 (0·0) | 0 (0·0) | 23 (19·2) | 0 (0·0) |
| Myositis | 0 (0·0) | 1 (1·6) | 0 (0·0) | 0 (0·0) | 0 (0·0) | 0 (0·0) | 0 (0·0) | 0 (0·0) | 0 (0·0) | 0 (0·0) | 0 (0·0) | 0 (0·0) |
| Oral candidiasis | 0 (0·0) | 0 (0·0) | 0 (0·0) | 0 (0·0) | 1 (0·8) | 0 (0·0) | 0 (0·0) | 1 (1·7) | 0 (0·0) | 0 (0·0) | 2 (1·7) | 0 (0·0) |
| Otitis media acute | 0 (0·0) | 8 (12·9) | 0 (0·0) | 0 (0·0) | 16 (13·4) | 0 (0·0) | 0 (0·0) | 9 (15) | 0 (0·0) | 0 (0·0) | 23 (19·2) | 0 (0·0) |
| Parasitic gastroenteritis | 0 (0·0) | 0 (0·0) | 0 (0·0) | 0 (0·0) | 1 (0·8) | 0 (0·0) | 0 (0·0) | 0 (0·0) | 0 (0·0) | 0 (0·0) | 3 (2·5) | 0 (0·0) |
| Parotitis | 0 (0·0) | 0 (0·0) | 0 (0·0) | 0 (0·0) | 1 (0·8) | 0 (0·0) | 0 (0·0) | 0 (0·0) | 0 (0·0) | 0 (0·0) | 0 (0·0) | 0 (0·0) |
| Pharyngitis | 0 (0·0) | 7 (11·3) | 0 (0·0) | 0 (0·0) | 5 (4·2) | 0 (0·0) | 0 (0·0) | 1 (1·7) | 0 (0·0) | 0 (0·0) | 9 (7·5) | 0 (0·0) |
| Poisoning | 0 (0·0) | 0 (0·0) | 0 (0·0) | 0 (0·0) | 0 (0·0) | 0 (0·0) | 0 (0·0) | 0 (0·0) | 0 (0·0) | 0 (0·0) | 1 (0·8) | 0 (0·0) |
| Prurigo | 0 (0·0) | 0 (0·0) | 0 (0·0) | 0 (0·0) | 0 (0·0) | 0 (0·0) | 0 (0·0) | 4 (6·7) | 0 (0·0) | 0 (0·0) | 12 (10) | 0 (0·0) |

| Arm | Delayed Rabies Control | | | Delayed RH5.1/Matrix-M | | | Monthly Rabies Control | | | Monthly RH5.1/Matrix-M | | |
|---|---|---|---|---|---|---|---|---|---|---|---|---|
| **Grade** | Mild | Mod. | Sev. | Mild | Mod. | Sev. | Mild | Mod. | Sev. | Mild | Mod. | Sev. |
| **Number of children followed** | **62** | | | **119** | | | **60** | | | **120** | | |
| Pyoderma | 1 (1·6) | 5 (8·1) | 0 (0·0) | 0 (0·0) | 21 (17·6) | 0 (0·0) | 0 (0·0) | 13 (21·7) | 0 (0·0) | 0 (0·0) | 33 (27·5) | 0 (0·0) |
| Pyrexia | 0 (0·0) | 0 (0·0) | 0 (0·0) | 0 (0·0) | 1 (0·8) | 0 (0·0) | 0 (0·0) | 0 (0·0) | 0 (0·0) | 0 (0·0) | 0 (0·0) | 0 (0·0) |
| Rhinitis | 0 (0·0) | 28 (45·2) | 0 (0·0) | 0 (0·0) | 61 (51·3) | 0 (0·0) | 0 (0·0) | 9 (15) | 0 (0·0) | 0 (0·0) | 41 (34·2) | 0 (0·0) |
| Skin candida | 0 (0·0) | 0 (0·0) | 0 (0·0) | 0 (0·0) | 0 (0·0) | 0 (0·0) | 0 (0·0) | 1 (1·7) | 0 (0·0) | 0 (0·0) | 1 (0·8) | 0 (0·0) |
| Staphylococcal skin infection | 0 (0·0) | 1 (1·6) | 0 (0·0) | 0 (0·0) | 0 (0·0) | 0 (0·0) | 0 (0·0) | 0 (0·0) | 0 (0·0) | 0 (0·0) | 0 (0·0) | 0 (0·0) |
| Thermal burn | 0 (0·0) | 0 (0·0) | 0 (0·0) | 0 (0·0) | 0 (0·0) | 0 (0·0) | 0 (0·0) | 0 (0·0) | 0 (0·0) | 0 (0·0) | 2 (1·7) | 0 (0·0) |
| Urinary tract infection | 0 (0·0) | 0 (0·0) | 0 (0·0) | 0 (0·0) | 1 (0·8) | 0 (0·0) | 0 (0·0) | 0 (0·0) | 0 (0·0) | 0 (0·0) | 0 (0·0) | 0 (0·0) |
| Wound | 0 (0·0) | 1 (1·6) | 0 (0·0) | 0 (0·0) | 0 (0·0) | 0 (0·0) | 0 (0·0) | 1 (1·7) | 0 (0·0) | 0 (0·0) | 1 (0·8) | 0 (0·0) |

#### 3.21 Table S20: Impact of vaccine arm on anaemia according to the secondary case definition (moderate).

Haemoglobin concentration <8 g/dL at 2 and 6 months post-third vaccine dose. *Fishers exact p (Freeman-Halton extension for 2x3 table) calculated when control cell=0. [+]Calculated on subset of children with complete data on ITN and SMC use at 2 months: Combined controls=99; Monthly RH5.1/MM =104; Delayed RH5.1/MM =102; and at 6 months: Combined controls=94; Monthly RH5.1/MM =104; Delayed RH5.1/MM =101.

| Group | N (%) | Odds ratio (95% CI) ; p [unadjusted] | Odds ratio (95% CI) ; p [adjusted[+]] |
|---|---|---|---|
| **2 MONTHS** | | | |
| **Combined control groups** | | | |
| Rabies controls (combined) n=105 | 1 (0·95) | 1 | 1 |
| Monthly RH5.1/MM n=113 | 4 (3·74) | 4·0 (0·44, 36·75) ; 0·2153 | 3·8 (0·41, 35·32) ; 0·2411 |
| Delayed RH5.1/MM n=107 | 3 (2·65) | 2·8 (0·29, 27·70) ; 0·3699 | 2·7 (0·27, 27·14) ; 0·3988 |
| **RH5.1/MM: delayed versus monthly** | | | |
| Monthly RH5.1/MM | 4 (3·74) | 1 | 1 |
| Delayed RH5.1/MM | 3 (2·65) | 0·70 (0·15, 3·21) ; 0·6488 | 0·76 (0·15, 3·92) ; 0·7417 |
| **6 MONTHS** | | | |
| **Combined control groups** | | | |
| Rabies controls (combined) n=100 | 0 (0·00) | Fisher's exact p = 0·1247* | - |
| Monthly RH5.1/MM n=112 | 2 (1·79) | | |
| Delayed RH5.1/MM n=103 | 4 (3·88) | | |
| **RH5.1/MM: delayed versus monthly** | | | |
| Monthly RH5.1/MM | 2 (1·79) | 1 | 1 |
| Delayed RH5.1/MM | 4 (3·88) | 0·45 (0·08, 2·51) ; 0·3626 | 0·52 (0·08, 3·49) ; 0·4984 |

#### 3.22 Table S21: Anti-RH5.1 serum IgG ELISA responses and percentage *in vitro* growth inhibition activity (GIA) of *P. falciparum* cultures by vaccination group.

Analyses at day 14 post-dose three (per-protocol sample). All available samples were assayed for ELISA: N=111 children for the Rabies (Rabivax-S) delayed and monthly groups combined, N=113 for the Delayed RH5.1/Matrix-M group, and N=107 for the Monthly RH5.1/Matrix-M group; and for GIA: N=97 children for the Rabies (Rabivax-S) delayed and monthly groups combined, N=113 children for the Delayed RH5.1/Matrix-M group, and N=108 for the Monthly RH5.1/Matrix-M group.

° Interquartile Range (IQR). *Effect size for anti-RH5.1 serum IgG responses: the ratio of geometric mean antibody concentration between groups estimated by linear regression of log10 responses. For GIA, the effect size is difference in mean percentage GIA between groups, with confidence intervals and p-values calculated by bootstrap. Effect size for % children with GIA greater than 60% is the odds ratio between groups estimated by logistic regression.

| Anti-RH5.1 serum IgG concentrations (µg/mL) by ELISA | | | |
|---|---|---|---|
| **Group** | **Geometric mean IgG concentration (IQR°)** | **Effect size* (95% CI)** | **p** |
| **Combined control groups v RH5.1/MM** | | | |
| Rabies controls (combined) | 0·03 (0·03, 0·03) | 1 | |
| Delayed RH5.1/MM | 836·60 (326·47, 2200·01) | 24648·37 (20988·24, 28946·80) | <0·0001 |
| Monthly RH5.1/MM | 626·49 (222·26, 1454·69) | 18457·94 (15682·22, 21724·95) | <0·0001 |
| **RH5.1/MM: Delayed versus Monthly** | | | |
| Monthly RH5.1/MM | 626·49 (222·26, 1454·69) | 1 | |
| Delayed RH5.1/MM | 836·60 (326·47, 2200·01) | 1·34 (1·13, 1·57) | 0·0006 |
| Percentage growth inhibition activity (GIA) of *P. falciparum* cultures by total purified IgG at 2·5 mg/mL | | | |
| **Group** | **Mean percent growth inhibition (SD)** | **Effect size* (95% CI)** | **p** |
| **Combined control groups v RH5.1** | | | |
| Rabies controls (combined) | 4·05 (12·20) | 0 | |
| Delayed RH5.1/MM | 79·02 (14·33) | 74·97 (71·37, 78·58) | <0·0001 |
| Monthly RH5.1/MM | 74·15 (15·88) | 70·10 (66·24, 73·97) | <0·0001 |
| **RH5.1/MM: Delayed versus Monthly** | | | |
| Monthly RH5.1/MM | 74·15 (15·88) | 0 | |
| Delayed RH5.1/MM | 79·02 (14·33) | 4·87 (0·91, 8·82) | 0·0159 |

| **Group** | **% children GIA >60% (n)** | **Effect size* (95% CI)** | **p** |
|---|---|---|---|
| **Combined control groups v RH5.1/MM** | | | |
| Rabies controls (combined) | 2·1% (2) | 1 | |
| Delayed RH5.1/MM | 87·6% (99) | 335·89 (74·34, 1517·61) | <0·0001 |
| Monthly RH5.1/MM | 81·5% (88) | 209·00 (47·47, 920·14) | <0·0001 |
| **RH5.1/MM: Delayed versus Monthly** | | | |
| Monthly RH5.1/MM | 81·5% (88) | 1 | |
| Delayed RH5.1/MM | 87·6% (99) | 1·61 (0·77, 3·37) | 0·2094 |
